# Spinal Cord Stimulation Improves Deceleration Phase Control during Targeted Reaching Post-Stroke

**DOI:** 10.1101/2025.10.29.25338778

**Authors:** Omar Refy, Luigi Borda, Jacob Hsu, Erick Carranza, Nikhil Verma, Roberto de Freitas, Erynn Sorensen, Amy Boos, Marc Powell, Lee Fisher, Peter Gerszten, Elvira Pirodini, John W. Krakauer, George F. Wittenberg, Hartmut Geyer, Marco Capogrosso, Douglas J. Weber

**Affiliations:** NeuroMechatronics Lab, Carnegie Mellon University, Pittsburgh, PA, USA; Physics Department, Carnegie Mellon University, Pittsburgh, PA, USA; Legged Systems Lab, Carnegie Mellon University, Pittsburgh, PA, USA; Mechanical Engineering Department, Carnegie Mellon University, Pittsburgh, PA, USA; Neuroscience Institute, Carnegie Mellon University, Pittsburgh, PA, USA; Robotics Institute, Carnegie Mellon University, Pittsburgh, PA, USA; Rehabilitation and Neural Engineering Labs (RNEL), University of Pittsburgh, Pittsburgh, USA; Reach Neuro, Inc., University of Pittsburgh, Pittsburgh, USA; Department of Physical and Rehabilitation Medicine, University of Pittsburgh, Pittsburgh, USA; GRECC, HERL, VA Pittsburgh Healthcare System and Department of Neurology, University of Pittsburgh, Pittsburgh, USA; Department of Neurosurgery, University of Pittsburgh, Pittsburgh, USA; Department of Neurology, Johns Hopkins University, Baltimore, USA; Department of Neuroscience, Johns Hopkins University, Baltimore, USA; Department of Physical Medicine and Rehabilitation, Johns Hopkins University, Baltimore, USA; Santa Fe Institute, Santa Fe, USA; Department of Neuroscience, University of Copenhagen, Copenhagen, Denmark

## Abstract

Cervical spinal cord stimulation (SCS) improves upper-limb function in individuals with chronic post-stroke hemiparesis, yet how it shapes motor control of arm movement remains unclear. During goal-directed reaching in healthy individuals, movements consist of a coordinated acceleration phase toward the target followed by a deceleration phase that stabilizes the limb near the endpoint. Disruptions in neuromotor control post-stroke can be partially restored by SCS, with prominent improvements occurring in the deceleration phase. To quantitatively characterize these effects, we used a proportional–derivative (PD) control model to fit planar reaching data from 12 healthy and 5 stroke participants. Movements were well described by the model, with proportional gain terms capturing the acceleration phase and derivative gain terms capturing velocity-dependent deceleration. In healthy individuals, model fits revealed a consistent balance between position and velocity-dependent torques that closely matched the optimal solution for smooth and stable reaching predicted by optimal feedback control simulations. In stroke, this balance was altered and partially normalized by SCS, with the most consistent changes observed in the velocity-dependent term. While the prevailing hypothesis is that SCS boosts motor drive to weak agonistic muscles, these results indicate a synergistic and potentially dominant effect in suppressing hyperexcitability of antagonistic muscles controlling the deceleration phase of movement. Finally, the PD controller model revealed frequency-dependent effects of SCS, suggesting that the model parameters may serve as biomarkers for guiding the selection of stimulation parameters. This work serves as a framework for characterizing how neuromodulatory therapies influence arm control .

## Introduction

Stroke is among the leading causes of disability worldwide^1^. As of 2019, stroke has a global prevalence of over 100 million people and has created a formidable health and economic burden, estimated at $2 trillion annually (∼1.7% of global GDP)^2,3^. In addition to the immense magnitude of this global problem, approximately 50% of stroke survivors experience permanent deficits in arm and hand function due to disruption of motor control circuits in the brain ^4^.

Recent studies demonstrate that neuromodulation of sensorimotor pathways in the spinal cord and brain may be effective for promoting recovery of motor function in individuals with chronic hemiparesis, even several years after stroke ^5–7^. By directly targeting neural circuits in the central nervous system (CNS), neuromodulation alters the dynamics of the neural circuits underlying the feedback control of movement. Thus, CNS neuromodulation directly promotes changes in the motor output of the CNS, which differs from traditional approaches that compensate for muscle weakness using robotic exoskeletons or functional electrical stimulation (FES) to assist movement (Fig. 1c). However, the direct effects of CNS neuromodulation in altering the dynamics of CNS motor control have yet to be examined.

**Fig. 1.**
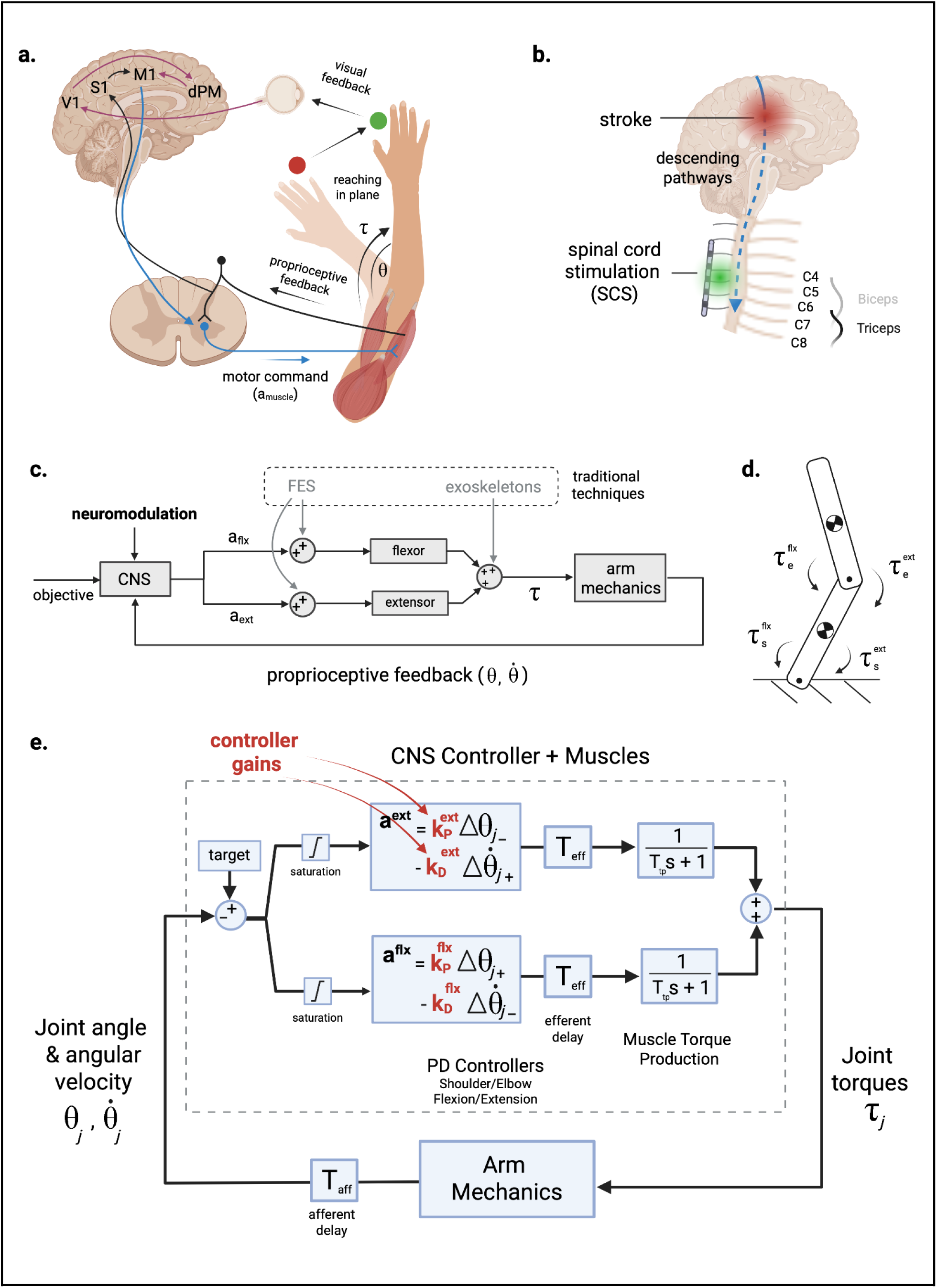
Modeling the feedback control of arm reaching and the effects of stroke and spinal cord stimulation. **a**, The neural control of arm movement spans the spinal cord and multiple brain regions to integrate visual and proprioceptive feedback of limb-state (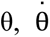) for controlling activation (**a**) of muscles that generate torques (τ) at each joint; figure adapted from Scott9. V1: primary visual cortex; S1: primary somatosensory cortex; M1: primary motor cortex; dPM: dorsal premotor cortex. **b**, Stroke disrupts neural circuits in the brain that communicate with the spinal cord, leading to severe motor impairments. Spinal cord stimulation (SCS) is a neuromodulation therapy that targets the primary sensory afferents innervating arm and hand muscles in the cervical spinal cord. **c**, Existing therapies, such as functional electrical stimulation (FES) and exoskeletons, attempt to compensate for the loss of motor output by stimulating the muscles or applying assistive torque at the joint, respectively. Neuromodulatory therapies (such as SCS) attempt to improve control of motor output from the spinal cord by targeting the central nervous system (CNS) directly. **d**, Schematic of two-link model used to model planar arm movements. **e**, The neural control of flexor and extensor muscle activation at each joint can be modeled using proportional-derivative (PD) controllers. Afferent signals encode limb-state information (joint angular position and velocity), which are delayed by nerve conduction times (T_aff_), while neural control outputs are delayed by T_eff_. Torque production at the muscle level is modeled as a first order transfer function of time constant T_tp_. Gaussian process based Bayesian Optimization is used to discover the neural controller gains (**k_P_** and **k_D_**) for each joint based on the joint kinematics and torques measured during point-to-point reaching movements.

Neuromotor control is achieved through a distributed network of feedforward and feedback circuits that span the brain and the spinal cord (Fig. 1a). Feedback control is a dominant mode of control for all types of motor behaviors, and there is strong evidence supporting a theory that feedback gains are optimized to generate movements that are smooth, energetically efficient, and stable in the presence of disturbances ^8–10^. Moreover, stroke is known to disrupt feedback control circuits, such as the stretch reflex, leading to excessive co-contraction of antagonistic muscles during reaching ^11,12^. Indeed, Borda et al.^13^ demonstrated recently that tonic SCS targeting elbow extensor muscles produced concurrent inhibition of reflex gains and motor unit activity in the elbow flexors. These changes were associated with improvements in reaching kinematics.

To understand the effects of SCS on the control of agonist-anatagonist muscle interactions during reaching, we created a computational model to simulate the mechanics and feedback control of joint torques at the shoulder and elbow. The arm was modeled as a two-link chain with pairs of uniarticular flexor and extensor muscles at the shoulder and elbow (see Fig. 1d). The activation of each muscle was modeled as an independent proportional-derivative (PD) controller with gain parameters (proportional gain: k_P_, derivative gain: k_D_) that were tuned to match the kinematics and torque profiles of human subjects performing planar, point-to-point reaching movements. Reaching data was collected from neurotypical (n=12) and hemiparetic participants (n=5) performing targeted reaching movements in a robotic exoskeleton (KINARM, Kingston, Ontario, Canada).

We found that the feedback gain parameters for neurotypical participants (n=12) were similar and matched those obtained by optimizing feedback gains to maximize trajectory smoothness in a dynamic simulation. The gain parameters differed significantly for reaching movements performed by people with stroke, with the largest differences appearing as increased derivative gains for the antagonistic muscles, which produce velocity-dependent torque that decelerates the joint during the terminal phase of reaching^14^. However, when SCS was applied, the controller gains changed to approximate those found for the healthy participants and optimal control simulations. The main effect of SCS was a strong and consistent reduction in the derivative gains across the arm joints, indicating improved control of velocity-dependent torque production at the terminal phase of the movement ^15^. Lastly, we found that the effects of SCS on controller gains varied with the SCS pulse-frequency, providing a mapping function that can be used to select SCS parameters that optimize the feedback control behavior of the CNS.

The experimental data and modeling results presented here imply that, rather than boosting motor drive at early phases of movement, SCS primarily enhances control of the deceleration and stabilization phase. In other words, the SCS-mediated improvements in reaching performance observed in this planar reaching task were due to a reduction in motor drive during the terminal phase of the movement, rather than an increase in motor drive during the acceleration phase. While these results do not diminish the importance of the SCS-mediated enhancements in agonist muscle activation, they reveal that SCS primarily enhances the neural controller performance, especially at the late phases of the movement. The model presented here provides a means to quantify changes in the arm control structure induced by SCS post-stroke, and can be used generally to understand the effect of neuromodulation therapies on different neurological diseases.

## Results

We start by quantifying the effects of SCS on reaching performance in five stroke survivors. We then present a PD controller model and show that it can simulate reaching behavior of both neurotypical and paretic arms. We show that in neurotypical participants, the PD controller gains exhibit a stereotypical structure, and that the controller gains for the paretic arm deviate from that pattern in a subject-specific fashion. Under SCS, the feedback controller gains are changed to become closer to neurotypical ranges in a stimulation-parameter dependent manner. Finally, we use optimal feedback control simulations to show that controller gains that are optimized to produce smooth point-to-point reaching movements match those found for neurotypical participants, suggesting that SCS tunes neural feedback controller gains towards optimal values.

### SCS Improves Planar Reaching of the Paretic Arm

We quantified the effects of cervical SCS on planar reaching in five individuals with chronic post-stroke hemiparesis. Participants performed a center-out planar reaching task under stimulation-off (STIM OFF) and stimulation-on (STIM ON) conditions. Performance was assessed using success rate, reach time, path efficiency, and smoothness quantified by the number of elbow-velocity peaks (Fig. 2; see Methods).

**Fig. 2.**
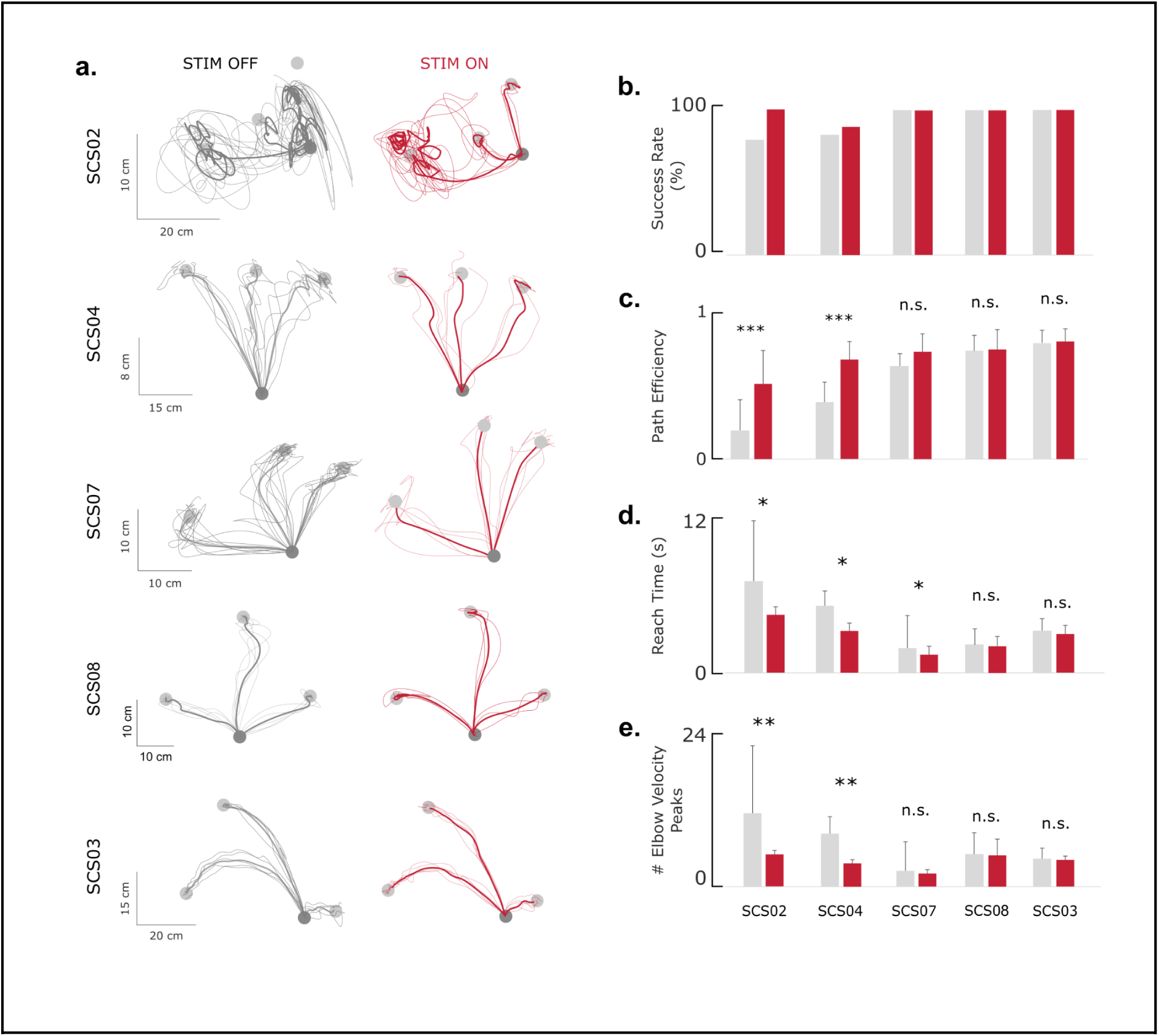
The effect of SCS on planar reaching in 5 stroke participants. **a**, Hand trajectories in a center-out task under STIM OFF and STIM ON conditions show varying degrees of impairment and improvement with SCS. **b**, Success rate of reaching the target under STIM OFF (gray) and STIM ON (colored). **c**, Path efficiency, defined as the ratio of the straight line path over the actual path length. **d**, Reach time, defined as the time taken to reach the target after the ‘go cue’ is given. **e**, Number of elbow velocity peaks as a measure of path smoothness. All but the least impaired participants (SCS03 and SCS08) showed a statistically significant improvement in at least one out of four metrics. Significance levels are indicated by * for p < 0.05, ** for p < 0.01 and *** for p < 0.001. Sample sizes are reported in Extended Data Table 2.

Across participants, SCS improved reaching kinematics, particularly in those with greater baseline impairment (Extended Data Table 1). The most impaired participant (SCS02) showed significant gains (p < 0.05) in all performance metrics. SCS04 exhibited significant improvements in path efficiency and smoothness, whereas SCS07 showed shorter reach times (p < 0.001). SCS03, who had mild impairment, showed no significant changes. Group-level averages (Fig. 2b–e) demonstrate consistent enhancements in success rate and path efficiency, reduced reach time, and fewer velocity peaks under SCS. These findings indicate that cervical SCS enhances movement fluency and stability during reaching, with effects that scale with baseline motor impairment.

### Feedback Control Model Reproduces Healthy and Paretic Reaching Behavior

To examine how SCS alters feedback control, we developed a proportional–derivative (PD) controller model that simulated joint torque control at the shoulder and elbow (Fig. 3; see Methods). The arm was modeled as a two-link chain, with flexor and extensor muscle controllers acting independently at each joint. Joint position and velocity feedback were delayed by 30 ms to represent afferent conduction, and controller outputs were delayed by 30 ms to simulate efferent transmission latencies. Muscle torque generation was modeled as a first-order process with a 50 ms time constant. Controller gains were tuned using Gaussian-process Bayesian optimization to minimize the error between simulated and experimental torques for individual trials.

**Fig. 3.**
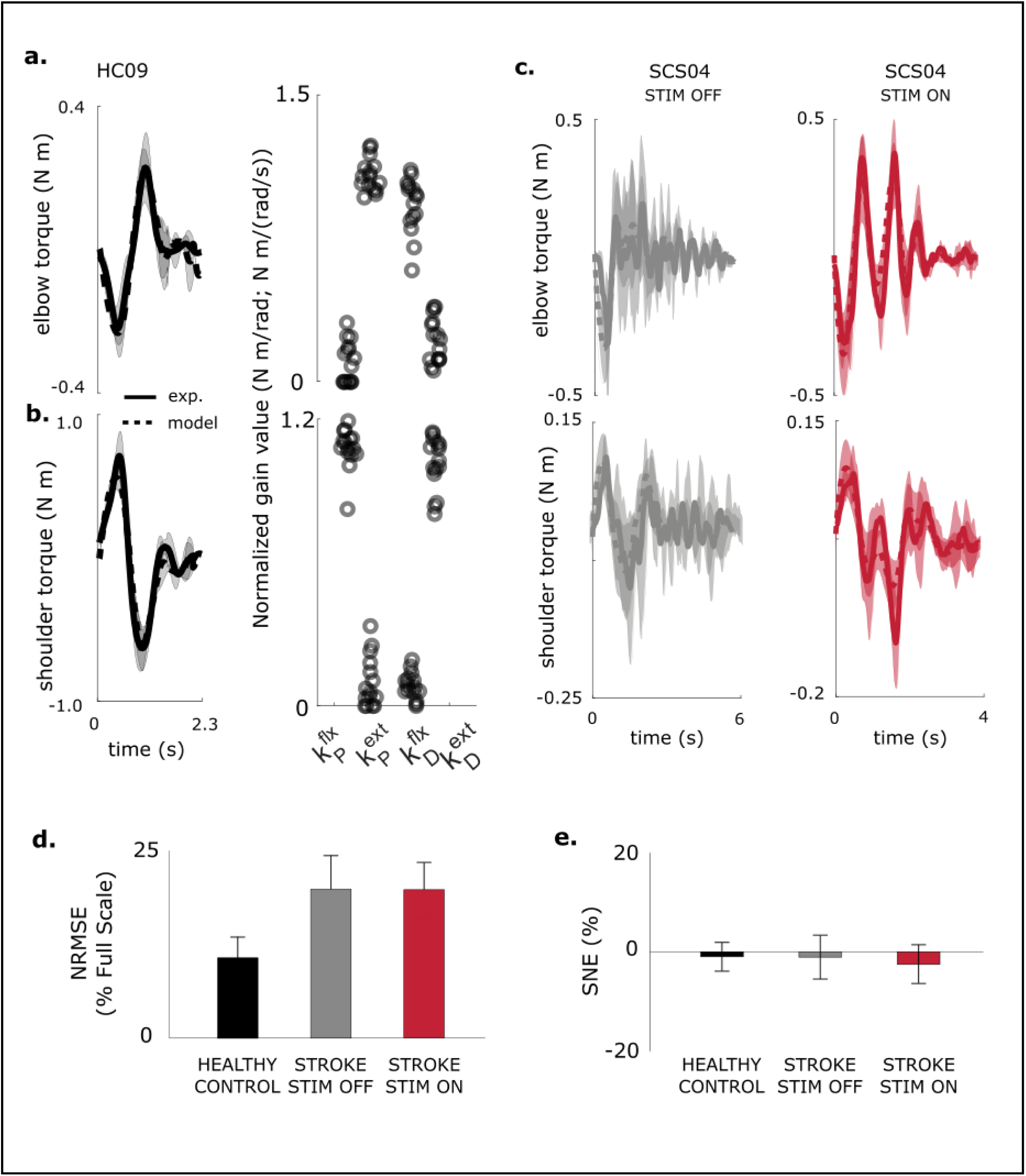
Model fitting performance across participants. **a-b**, Model fit of elbow and shoulder torque profiles for a neurotypical healthy control participant (HC09). Resulting model control gains followed a U-shape for shoulder (and inverted U-shape for elbow) for all control participants. Sample size is 5 repetitions for experimental data, and 25 model fits (5 per repetition). **c**, Example model fits for shoulder and elbow torque profiles under STIM OFF and STIM ON conditions (SCS04). **d-e**, Summary of model performance based on NRMSE and SNE across control and stroke participants under STIM OFF and STIM ON conditions. Across all participants, the model reproduced torque profiles with 86% accuracy (based on NRMSE), while tracking average torque with 99% accuracy (based on SNE). Sample size is reported in Extended Data Table 2.

In healthy participants, the model reproduced measured joint torques with high accuracy (average NRMSE = 10.7 ± 2.8%, SNE = –1.0 ± 2.8%). Torque traces and fitted gains showed characteristic U-shaped and inverted-U gain structures for the elbow and shoulder, respectively (Fig. 3a,b).

For the stroke group (n = 5; SCS02, SCS03, SCS04, SCS07, SCS08), the model also captured behavior across STIM OFF and STIM ON conditions with comparable accuracy (NRMSE = 18.9 ± 4.3% and 17.9 ± 5.1%; SNE = –0.7 ± 4.4% and –1.8 ± 3.6%, respectively; p > 0.3 for all comparisons). NRMSE was significantly lower for healthy participants (p < 0.001), reflecting greater trial-to-trial variability after stroke. Overall, the PD controller reproduced torque profiles with ≈ 86% accuracy across all participants and conditions, validating it as an effective model of feedback control of muscle activation in this point-to-point planar reaching task.

### PD controller gains

We next examined how feedback controller gains contribute to joint torque production during reaching. During extension movements, the proportional gain for the agonist (k ^ext^) and the derivative gain for the antagonist (k ^flx^) regulate the joint torques that accelerate and decelerate the joint, respectively during the initial and terminal phases of the reach (Fig. 4a; Supplementary Video 1). The remaining gains (k ^flx^, k ^ext^) contribute only small torques to stabilize the joint near the target. The fitted gains for the elbow muscles (Fig. 4b) form a consistent inverted-U pattern across all healthy control participants, reflecting the balance between movement initiation (“ballistic gains”) and termination (“stabilization gains”).

**Fig. 4.**
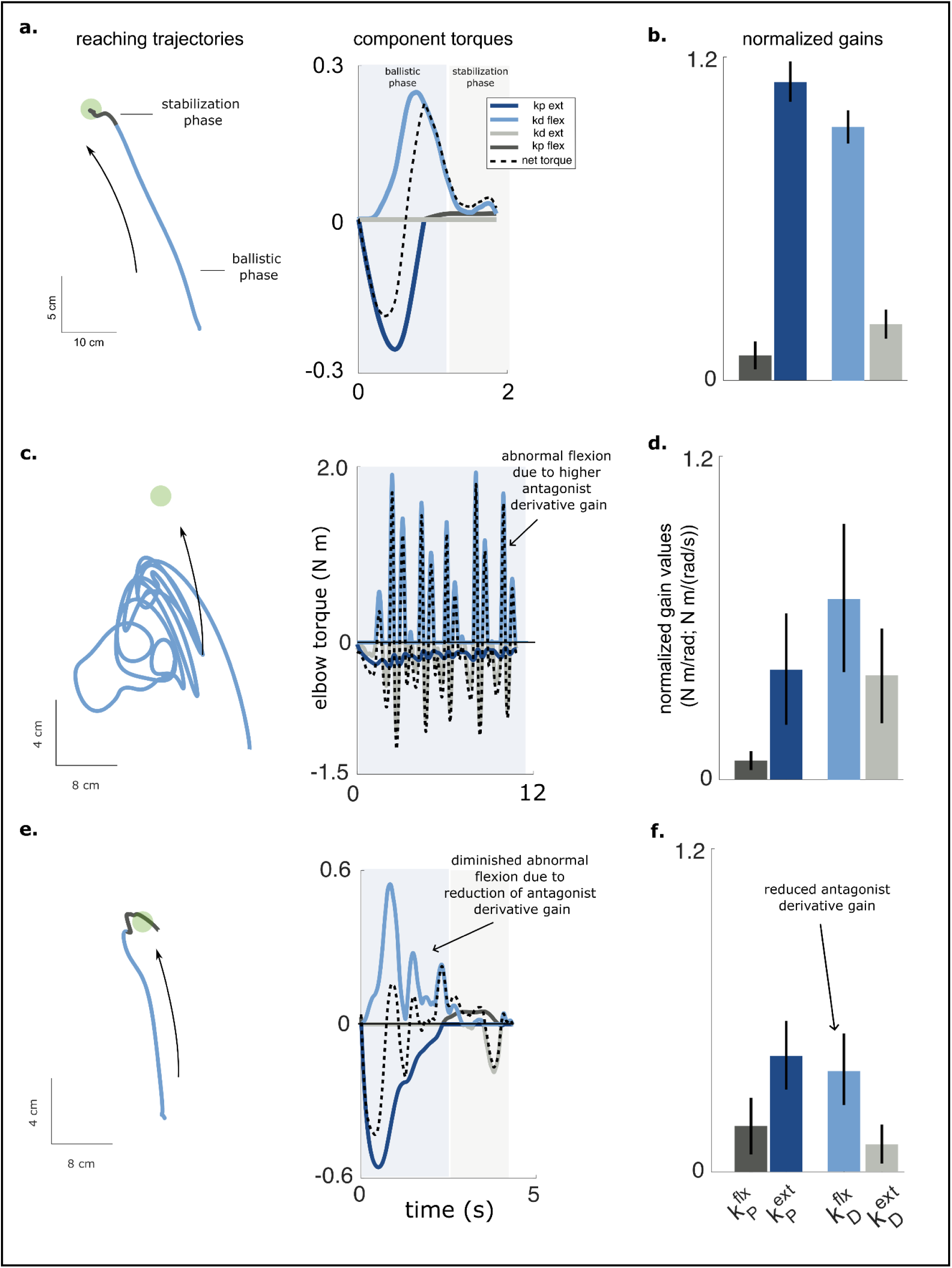
Feedback controller gains and their relative contributions to total torque output. **a**, Reaching trajectory of a control participant (HC09) and the associated component and total torque profiles for the elbow. **b**, The component torques are plotted in different colors and represent the contributions of each controller gain. The movement typically consists of a *ballistic phase* where the hand accelerates toward the target and then decelerates on approach to the target. The terminal or *stabilization* phase occurs at the end where fine corrections are made to stop at the target. **c**, Average controller gains for the elbow (HC09); the gains are normalized by their relative contribution to the total torque. We labeled the elbow extension proportional gain and flexion derivative gain as the ballistic gains (dark and light blue), and the flexion proportional and extension derivative gains as the stabilization gains (dark and light gray), reflecting their primary contributions to different phases of the movement. The ballistic gains dominate as the stabilization gains contribute only a small percentage of the total torque, resulting in an inverted U-shaped structure, which was observed across all healthy participants. **d**, Reaching trajectory of SCS02 under the STIM OFF condition shows erratic movements as the participant struggles to reach the target. **e**, The model reveals that the net elbow torque is dominated by large and alternating flexion and extension torques associated with the derivative gains. **f**, The structure of the controller gains for SCS02 differs from that of the healthy (control) participant. In this example, it is characterized by abnormally high elbow flexion and extension derivative gains. **g**, The reach trajectory for SCS02 improves markedly in the STIM ON condition. **h**, The profile of the component torques shows fewer oscillations and smaller peak torques when SCS is active. **i.** The derivative gains for both the flexor and extensor muscles are decreased closer to levels exhibited by the healthy control participant. Sample size for gain fits is 15 for both control and stroke participants (5 model fits per repetition, 3 repetitions).

In stroke participants, this gain structure was disrupted. For example, in SCS02 under the STIM OFF condition, large alternating flexion and extension torques prevented successful reaching (Fig. 4c; Supplementary Video 2). The model revealed abnormally high derivative gains for both flexors and extensors, producing excessive velocity-dependent torques that obstructed movement (Fig. 4d). With SCS, reaching performance improved (Fig. 4e; Supplementary Video 3), and derivative gains decreased toward healthy ranges (Fig. 4f), reducing abnormal oscillatory torques. Across participants (n = 5), SCS consistently shifted gain structures toward neurotypical patterns. These results suggest that the effects of stroke on neuromotor control are dominated by elevated derivative gains, resulting in excessive velocity-dependent torques, and these effects are mitigated by SCS to restore a more typical feedback control regime.

### Feedback Controller Gains Shift Toward Neurotypical Levels Under SCS

We next quantified how SCS affected the feedback controller gains obtained from model fits in each stroke participant (n = 5; Fig. 5). For brevity, we focus on the elbow joint during reaching to the central target; shoulder results are provided in the Supplementary Material. The bar plots show average values for the proportional and derivative gains for the flexor and extensor controllers under STIM OFF (grey) and STIM ON (red) conditions. The yellow shaded regions indicate the range of gain values found for each participant’s nonparetic limb performing the same reaching task.

**Fig. 5.**
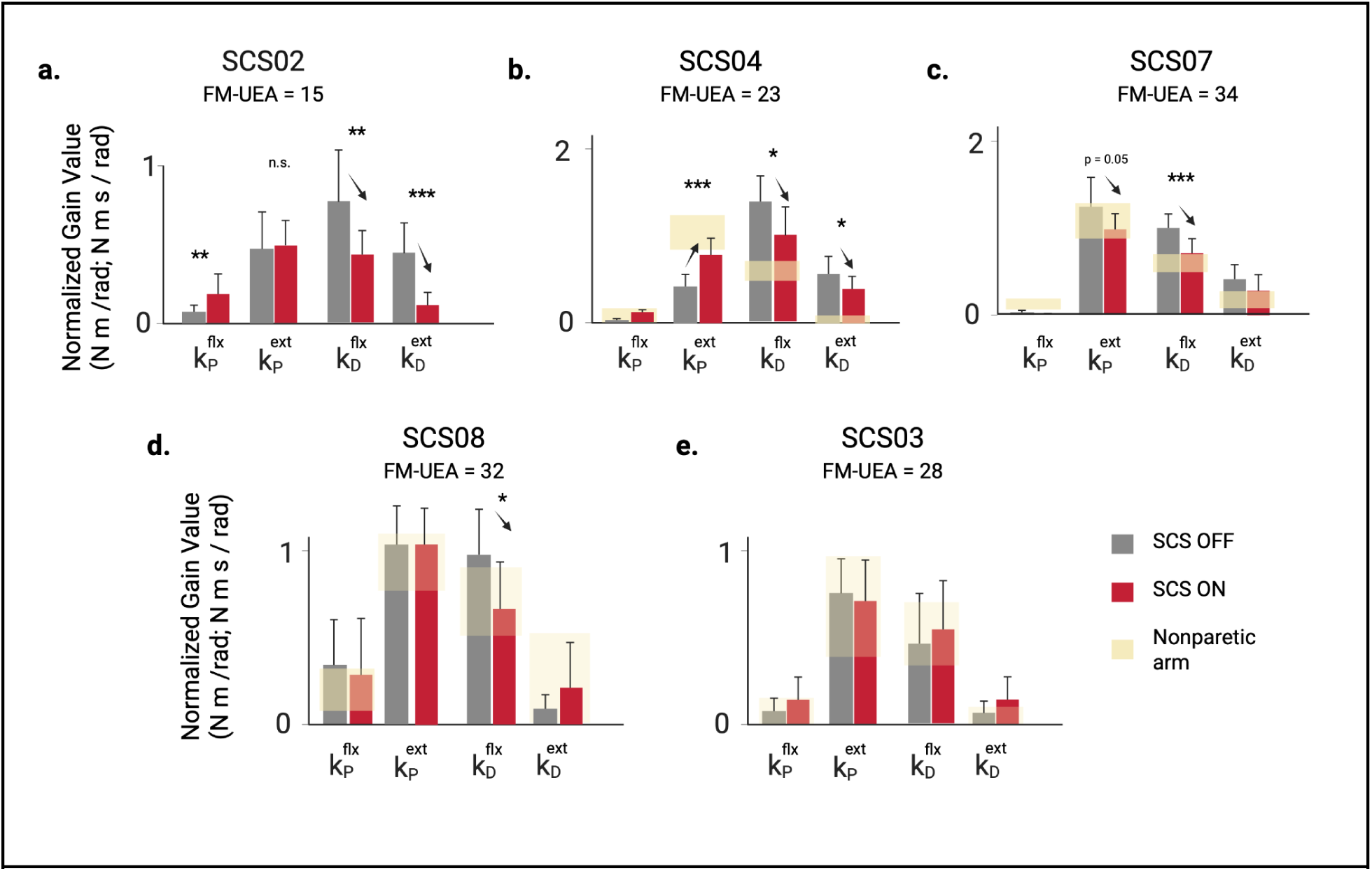
Feedback controller gains from model fitting to experimental data collected during the STIM OFF and STIM ON conditions. Bar plots showing the average gain values taken across 3-10 repetitions of the planar reaching task to a single target in each condition (model optimizations per repetition; individual repetition count per participant reported in Extended Data Table 3). The error bars indicate one standard deviation. The yellow shading indicates the range of gain values found for the nonparetic arm in participants SCS03-SCS08; planar reaching data was not collected for the nonparetic limb with SCS02. With the exception of SCS02, all participants showed significant reductions in the flexor derivative gain with STIM ON, while increases in the extensor proportional gain were observed in only one participant. Statistical significance is reported as t-tests (corrected for multiple comparisons; FDR Benjamini-Hochberg) between STIM OFF and STIM ON. Statistical significance: *, ** and *** denote p < 0.05, p < 0.01 and p < 0.001, respectively.

The most impaired participant (SCS02), showed the strongest tuning effects as the flexor derivative gain decreased by 44% (*p* = 0.0021, *d* = 1.19) and extensor derivative gain decreased by 73% (*p* < 0.001, *d* = 1.51), both shifting toward healthy ranges. Interestingly, the extensor proportional gain was unchanged by stimulation, indicating that SCS did not increase the elbow extension torque even though stimulation was targeting the triceps muscle. SCS04 also exhibited significant reductions in derivative gains (–26% and –30%, *p* < 0.05). Unlike SCS02, these were accompanied by an 83% increase in the extensor proportional gain (*p* < 0.001), reflecting the direct effect of SCS targeting the triceps to boost voluntary recruitment. SCS07 showed a 31% reduction in flexor derivative gain (*p* < 0.001) and a 21% decrease in extensor proportional gain (*p* = 0.05). SCS08 exhibited a 32% reduction in the flexor derivative gain (*p* = 0.031), bringing it into the range observed for their nonparetic arm. SCS03 displayed no significant changes, likely because their baseline reaching performance and controller gains were already within range of their nonparetic limb.

Across participants, the most consistent effect of SCS was a reduction in excessive derivative gains (k_D_), reflecting normalization of the level of velocity-dependent torque produced to slow the joint during the terminal phase of the reaching movement. Surprisingly, we observed only one case where the proportional gain (k_P_) for the extensor muscle increased significantly. Indeed, SCS04 was the only participant that exhibited a low proportional gain in the extensor muscle at baseline, and SCS boosted the gain to a level just below the nonparetic range. This pattern of changes contradicts the idea that SCS enhances voluntary recruitment of targeted muscles, such as triceps in the present analysis. Instead, these results suggest the dominant effect of SCS, used here to target triceps, is actually more effective in suppressing velocity-dependent torques generated by the biceps.

### Tuning of Model Gains Varies with Stimulation Frequency

When used clinically, SCS and nearly all other neuromodulation devices are tuned manually, often without a clear biomarker for measuring a therapeutic effect^16^. Since the patient’s response to neuromodulation depends critically on the choice of stimulation parameters, we used the feedback controller model to examine how the proportional and derivative gains change as stimulation frequency is adjusted to optimize reaching performance in two participants (Fig. 6). For SCS02, increasing SCS frequency from 0 Hz to 100 Hz progressively improved movement quality (Fig. 6a–e). Reach trajectories became straighter and smoother, exhibiting incremental improvements with each step in SCS frequency (*p* < 0.05). Fitted controller gains revealed that both flexor and extensor derivative gains declined with frequency, driving the overall gain structure toward the healthy pattern (Fig. 6b,e). These results demonstrate a stimulation-frequency dependent effect in reducing the flexor derivative gain at the elbow.

**Fig. 6.**
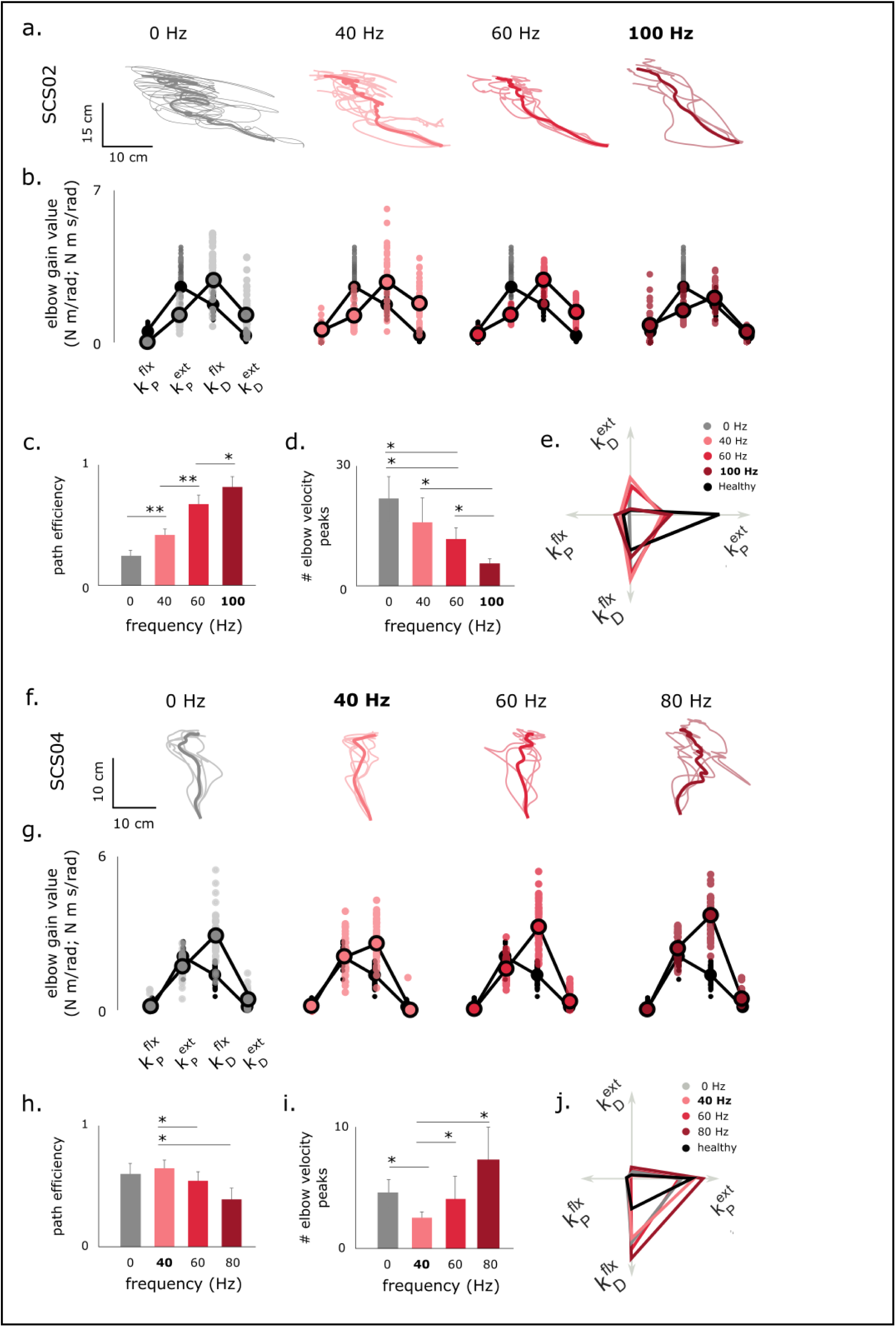
The effects of neuromodulation on reaching performance vary with SCS frequency. **a**, Reaching trajectories of SCS02 during free forward reaching under STIM OFF and STIM ON at 40, 60 and 100 Hz. Note the improvement in performance as stimulation frequency increases. **b**, Model normalized gains of the elbow joint in comparison to healthy ranges. Healthy ranges are indicated in transparent gray on all plots. Notice the restoration of the inverted U-shaped model gains as stimulation frequency increases. **c** and **d** show path efficiency and number of elbow velocity peaks at different stimulation frequencies. 100 Hz stimulation frequency resulted in straighter and smoother reaching paths. Sample size is 5 repetitions per frequency. **e.** Radar plots for average gain values. Notice how derivative gains are gradually reduced closer to healthy ranges. Panels **f-j** show the same results but for SCS04, where the optimal stimulation frequency was 40 Hz. For SCS04, increasing the stimulation frequency decreased reaching performance. Sample sizes for panels h and i are 5 repetitions for 0, 40 and 80 Hz and 10 repetitions for 60 Hz. Optimal frequencies are indicated in bold throughout the figure. Statistical significance is indicated as * for p < 0.05 and ** for p < 0.01.

A different trend was observed with SCS04 (Fig. 6f–j). The best performance occurred with an SCS frequency of 40 Hz, where reaches were smoothest and closest to healthy behavior. Path efficiency decreased from 0.66 at 40 Hz to 0.55 (*p* < 0.05, *d* = 1.20) and 0.39 (*p* < 0.05, *d* = 1.68) at 60 Hz and 80 Hz, respectively, while the number of velocity peaks increased from 2.6 to 4.2 and 7.4 (*p* < 0.05). At 80 Hz, both metrics were worse than baseline (STIM OFF; *p* < 0.05). Correspondingly, derivative gains diverged from healthy ranges as frequency increased (Fig. 6g,j).

Together, these results demonstrate that stimulation frequency strongly modulates reaching performance and the feedback controller model identified consistent reductions in the flexor derivative gain as a key contributor to the improvement in reaching behavior. While optimal tuning (e.g., 100 Hz for SCS02, 40 Hz for SCS04) enhances smooth, stable reaching, suboptimal frequencies can degrade performance, underscoring the need for individualized parameter optimization.

### Optimal Feedback Control Model Explains Gain Tuning

Because healthy participants exhibited a stereotypical U-shaped gain structure, we hypothesized that these feedback gains are tuned to optimize movement smoothness during targeted reaching. To test this, we simulated a two-link arm controlled by unidirectional flexor and extensor torque generators at the shoulder and elbow (Fig. 7; see Methods). Bayesian optimization was used to identify the proportional and derivative gain values that maximized trajectory smoothness while reaching a target within a specified duration (2 s).

**Fig. 7.**
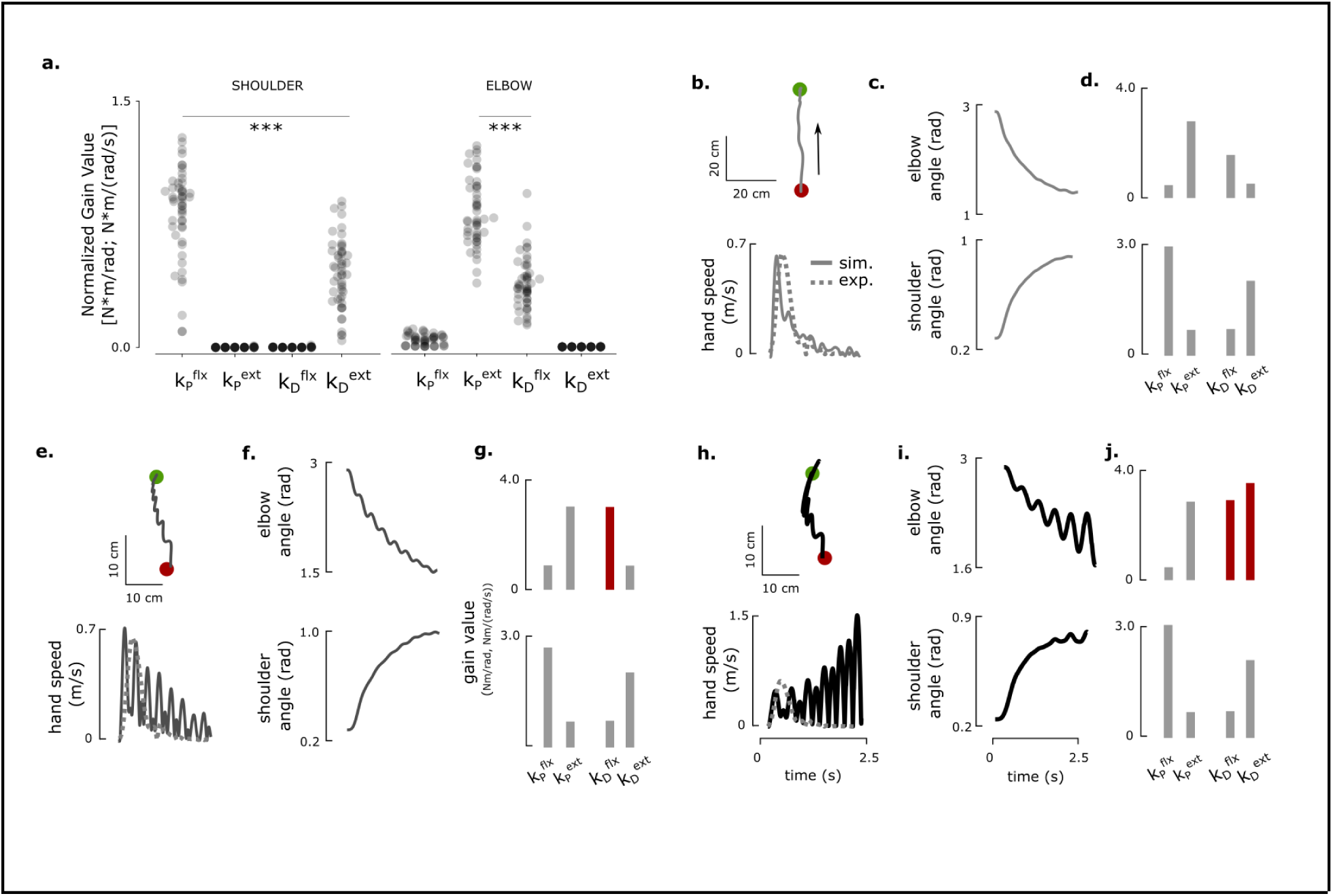
Optimal feedback control model reveals a set of feedback controller gains that match the pattern of those observed in neurotypical participants and explains deviations from optimal. **a**, The optimal elbow and shoulder gain structures resulting from optimizing a simulated arm’s trajectory smoothness time to target. The gains are normalized by their relative contributions to the total torque (as in Figure 4, panels c, f and i, Figures 5 and 6). The U-shaped and inverted U-shaped gain structures come out naturally from model optimization with an objective function to reach the target successfully with minimum jerk, where the ballistic gains generate the majority of torque. **b-j** show model reaching trajectories and joint angles using optimal gain structures (panel d), mistuned (50% increase) elbow flexor derivative gain (panel g) and mistuned elbow flexor (50% increase) and extensor derivative (700% increase) gains (panel j). Shoulder gains were kept at optimal ranges throughout. Mistuning elbow flexor derivative gain results in jerky reaching trajectories (panels **e-f**). If elbow extensor derivative gain is also mistuned, elbow movements (and hence shoulder through interaction torques) can become unstable (panels **h-i**). Dashed lines in panels h, k and n represent example data from healthy participant HC09. *** indicates p < 0.001.

Optimization consistently yielded the same gain organization observed experimentally: a U-shaped pattern at the elbow and an inverted U-shape at the shoulder (Fig. 7a; Supplementary Video 4). The extensor proportional and flexor derivative gains dominated torque production at the elbow, while the flexor proportional and extensor derivative gains dominated at the shoulder, matching the empirical findings in healthy participants.

Simulations using these optimal gains produced straight hand paths, bell-shaped velocity profiles, and smooth joint trajectories (Fig. 7b–d). Increasing the elbow flexor derivative gain alone induced mild oscillations in hand velocity (Fig. 7e–g; Supplementary Video 5), whereas simultaneously increasing both flexor and extensor derivative gains produced large, alternating flexion–extension torques (Fig. 7h–j), resembling behaviors observed in stroke participants such as SCS02 and SCS04.

These results demonstrate that the stereotypical gain structure observed in healthy participants corresponds to an optimal feedback control policy for generating smooth, stable reaching. Deviations from this structure result in clear and characteristic motor deficits, reinforcing that post-stroke impairments—and their restoration by SCS—can be understood as shifts in the multidimensional space of feedback control gains.

## Discussion

For decades, spinal cord stimulation (SCS) has been an established treatment for chronic pain. More recently, its potential to promote motor recovery after spinal cord injury^17–19^ and stroke^20^ has emerged as a major new application of neuromodulation. With ongoing efforts to translate SCS into neurorehabilitation practice, it is crucial to understand its mechanisms of action^21^, articulate its effect on motor control of voluntary movement, and develop principled approaches for programming stimulation parameters to maximize functional benefit.

When SCS is applied near the dorsal root entry zone, it primarily activates large-diameter afferent neurons, particularly Ia muscle afferents^22^. These afferents project directly and indirectly to spinal motor neurons, but also to many sensorimotor areas of the nervous system. At higher amplitudes, SCS can evoke trans-synaptic activation of motor neurons, evoking the posterior root muscle (PRM) reflex^23^. In neuromotor rehabilitation, however, SCS is applied below the motor threshold and delivered continuously at a fixed frequency, modulating excitability rather than directly eliciting contractions. Continuous activation of Ia afferents can thus increase motoneuron responsiveness to descending inputs^24,25^, potentially compensating for reduced corticospinal drive after stroke. In this context, SCS does not directly generate movement but modifies the input–output properties of the spinal (and CNS) circuitry, effectively tuning feedback gains within corticospinal control loops.

Here, we tested the hypothesis that SCS improves motor control by boosting motor drive during the acceleration (i.e. ballistic) phase of movement. We used a neuromechanical model to parse out position and velocity-dependent components of the torques generated at the shoulder and elbow during the acceleration and deceleration phases of reaching, respectively. In neurotypical (i.e. healthy) participants, we found that these component torques and associated controller gains exhibit stereotypical patterns (Figs. 3–4, Extended Data Figs. 1–2, 5) that are optimal for smooth, stable point-to-point reaching (Fig. 7). Stroke disrupted this gain structure (Figs. 4–5, Extended Data Figs. 3–4), but SCS partially restored it in a frequency-dependent manner (Fig. 6). We found that, rather than simply boosting agonist muscle drive, SCS suppresses velocity-dependent torques generated by antagonistic muscles during the deceleration phase of reaching. Thus, SCS enhances *control* of muscle activation more globally to achieve efficient regulation of joint torques throughout all phases of movement.

By decomposing joint torques into position- and velocity-dependent components, the model reveals how different feedback elements contribute to impaired movement and recovery. When SCS was active, the derivative gains, which generate velocity-dependent torques, decreased by 26–44% (Extended Data Fig. 3). The effect of SCS on proportional gains, while present in some participants, was far less consistent. However, the planar reaching task used here did not reveal clear deficits during the acceleration phase, which is when the position-dependent torques are dominant. Indeed, more challenging tasks, such as unsupported 3D reaching^5^, are more likely to reveal improvements in the proportional gains with SCS.

As shown in Fig. 6, optimal frequencies varied across participants and electrodes: in SCS02, higher frequencies (100 Hz) improved trajectory smoothness and gain normalization, whereas in SCS04, optimal performance occurred at lower frequencies (40 Hz). At suboptimal frequencies, performance deteriorated, emphasizing that SCS can both enhance or impair control depending on tuning. These results underscore the need for personalized optimization of stimulation parameters and suggest that models such as ours could help guide parameter selection by targeting specific gain profiles.

Like all models, ours has several limitations. It is important to note that our model attempts only to *describe* the behavior of the CNS in controlling joint torques during goal-directed reaching, and it is not intended to implicate specific neural circuits. The model captures only 86% of NRMSE variance, which leaves room for improvement, perhaps by including unmodeled factors such as changes in feedforward drive or multi-joint coordination deficits. Furthermore, the reaching behavior studied here is restricted to planar reaching without the influence of gravity.

Despite its simplicity, the model presented here provides an objective, quantitative method to assess changes in control at different phases of the movement induced by SCS. Through this approach, it becomes evident that healthy reaching is generated via optimal gain patterns that are optimal for smooth, stable movements, that stroke disrupts this pattern, and that SCS can partially restore it primarily by normalizing derivative gains that stabilize motion rather than by amplifying proportional gains that drive it. In other words, SCS enables the CNS to solve the control problem of reaching more efficiently. These findings bridge motor control theory and neurorehabilitation, offering a principled foundation for developing individualized, model-based interventions to restore movement after neurological injury.

## Methods

### Participants

This study was conducted as a substudy under the *Spinal Cord Stimulation for Restoration of Arm and Hand Function in People With Subcortical Stroke* (NCT04512690) trial aimed at evaluating the effects of spinal cord stimulation for restoring arm and hand function post-stroke. A total of seven participants completed the parent study, and five of the seven were selected for the experiments presented here. A group of 12 neurologically intact participants were tested (healthy controls, HC group). The five stroke participants (2M/3F) were 3-10 years post-stroke and whose impairment levels spanned a representative range of Fugl-Meyer scores. Participants were implanted with epidural spinal cord stimulation (SCS) electrodes for 29 days, which were removed following completion of the study. All experiments reported in this work were performed during the implant period. Participants in the stroke groups are referred to as SCSXX and are numbered based on their study enrollment order. Participant numbering is consistent with Powell et al.^20^ In addition, twelve healthy control participants (referred to as HCXX) with no underlying neurological disorders relevant to the upper extremity were recruited. Participant details are provided in Table 1. All experiments were approved by the University of Pittsburgh Institutional Review Board, and were conducted in accordance with the Declaration of Helsinki. All participants provided written informed consent.

**Table 1:**
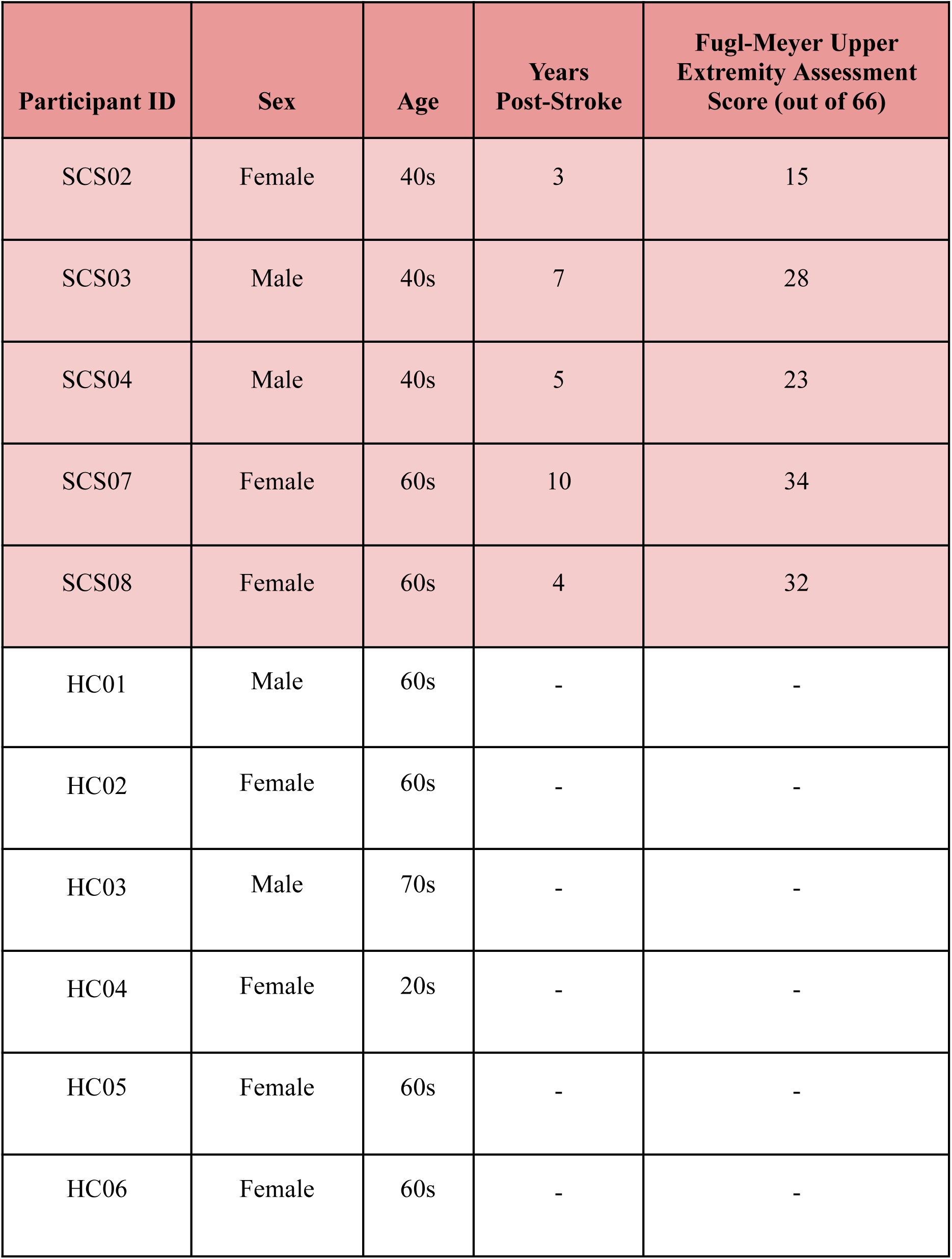
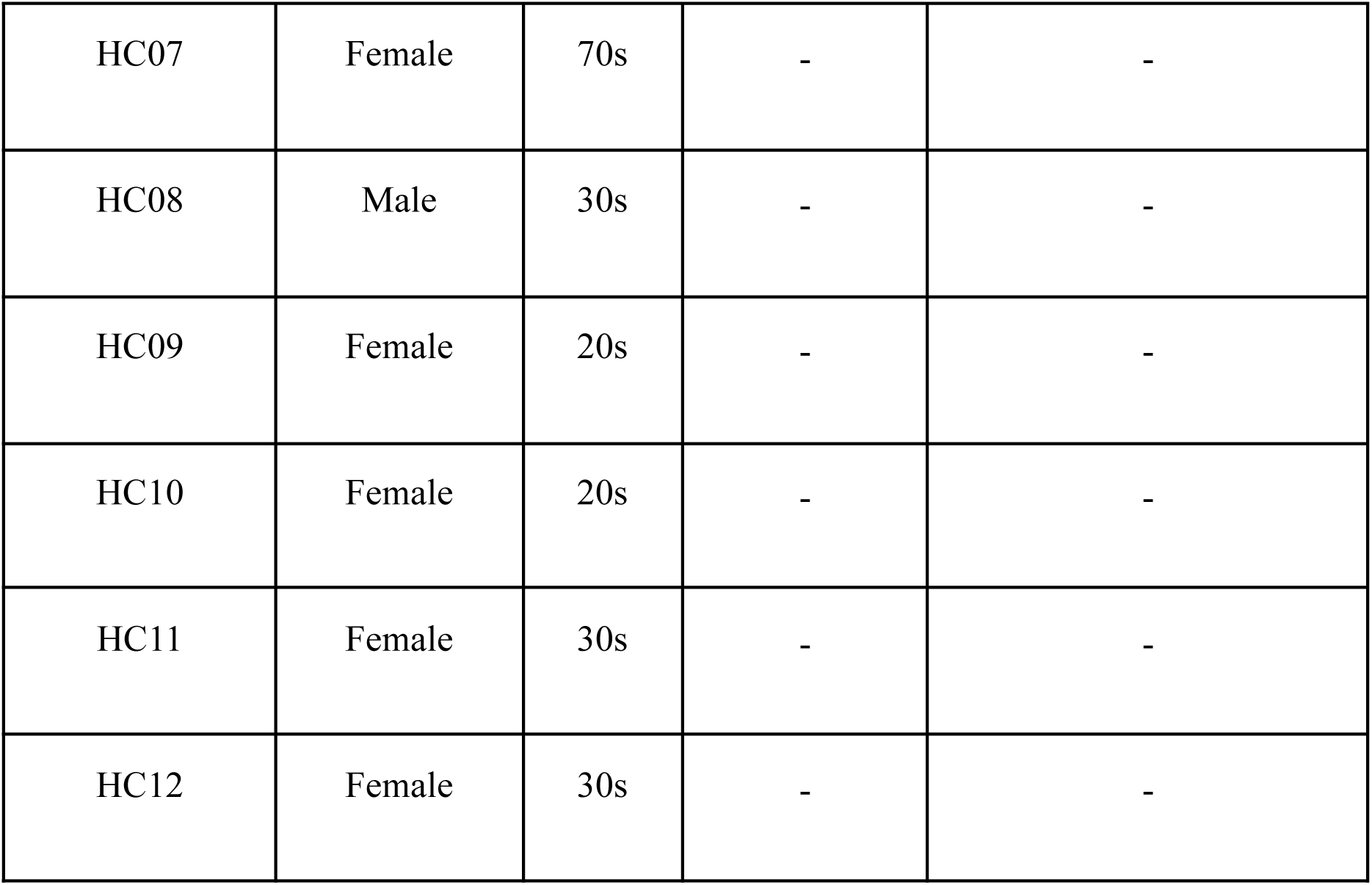
Study participant information.

| Participant ID | Sex | Age | Years Post-Stroke | Fugl-Meyer Upper Extremity Assessment Score (out of 66) |
| --- | --- | --- | --- | --- |
| SCS02 | Female | 40s | 3 | 15 |
| SCS03 | Male | 40s | 7 | 28 |
| SCS04 | Male | 40s | 5 | 23 |
| SCS07 | Female | 60s | 10 | 34 |
| SCS08 | Female | 60s | 4 | 32 |
| HC01 | Male | 60s | - | - |
| HC02 | Female | 60s | - | - |
| HC03 | Male | 70s | - | - |
| HC04 | Female | 20s | - | - |
| HC05 | Female | 60s | - | - |
| HC06 | Female | 60s | - | - |
| HC07 | Female | 70s | - | - |
| HC08 | Male | 30s | - | - |
| HC09 | Female | 20s | - | - |
| HC10 | Female | 20s | - | - |
| HC11 | Female | 30s | - | - |
| HC12 | Female | 30s | - | - |

### Implant and Intraoperative Testing

Stroke participants were implanted with two linear, 8-channel SCS leads (referred to as rostral and caudal, or R and C throughout this study) that were inserted into the epidural space of the cervical spinal cord to target the dorsal root entry zone at the spinal segments innervating the upper limb. Implant procedure details are described in Powell et al.^20^

Recruitment curves were measured intraoperatively and on the first day of testing to measure muscle activation thresholds for each SCS electrode. Briefly, stimulation pulses were applied at a low repetition rate (0.5-2 Hz), and evoked responses were measured in major muscle groups of the arm while stimulation amplitude was increased incrementally. The peak-to-peak amplitude of the compound action potential evoked in each muscle was used to quantify the strength of recruitment for each of the SCS electrodes, which were distributed along the rostro-caudal axis.

The recruitment thresholds for each muscle were used to guide the selection of stimulation parameters for targeting specific muscles. During the reaching experiments studied here, stimulation was applied at a fixed amplitude and frequency on one or more SCS electrodes, with parameters adjusted manually to optimize task performance. The stimulation amplitude was set to a level below the motor threshold found during the recruitment curve measurements, and the stimulation frequency was set to a level between 40 and 100 Hz. The parameter configurations used in the experiments reported in this study are summarized in Table 2.

**Table 2.** Stimulation parameters for all experimental trials used in the study. R and C stand for rostral and caudal, respectively, and refer to the rostral and caudal positions of the linear probes, respectively. The number preceding the R and C designation refer to the stimulation site (1-8) on the linear probes. The asterisk (*) indicates the best parameter set used in the SCS frequency optimization experiment shown in Figure 6.

| **Subject** | **Figures** | **Stimulation Parameters** [contact amplitude frequency] |
| --- | --- | --- |
| SCS02 | 6 | 5C 4.6mA 40Hz |
| SCS02 | 6 | 5C 4.6mA 60Hz |
| SCS02 | 2,4,5,6 | *5C 4.6mA 100Hz |
| SCS03 | 2,5 | 1R 1.4mA 60Hz, 4R 2.8mA 60Hz, 7R 2.0mA 60Hz |
| SCS04 | 2,3,5,6 | *5R 3.6mA 40Hz, 8R 3.9mA 40Hz |
| SCS04 | 6 | 5R 3.6mA 60Hz, 8R 3.9mA 60Hz |
| SCS04 | 6 | 5R 3.6mA 80Hz, 8R 3.9mA 80Hz |
| SCS07 | 2,5 | 4R 1.4mA 40Hz |
| SCS08 | 2,5 | 7R 2.6mA 60Hz |

**Table 3.** Inertial properties for estimating experimental torque and for the two-link model simulations.

|  | Upper Arm | Forearm |
| --- | --- | --- |
| Mass (kg) | 2.8 | 2.1 |
| Length (m) | .35 | .35 |

### 2D reaching tasks and data collection

During the 29 day implant period, the participants performed a variety of tasks such as 2D and 3D reaching tasks, and other functional assessments such as the Fugl-Meyer Upper Extremity Assessment (FM-UEA). Here, we focus only on the planar reaching task, which was performed using the Kinarm robotic exoskeleton (BKIN Technologies, Ltd., Kingston, Ontario, Canada) that supported the arm against gravity and measured kinematics at the shoulder and elbow at a 1000 Hz sampling rate. Joint torques were calculated using measured joint kinematics (see subsequent subsections for more details).

Participants were prompted to reach forward from a proximal starting position, near the chest, to targets positioned ∼8-40 cm away. The target locations were tailored to each participant’s range of motion and impairment level. The task was modified for participant SCS02, as they were unable to perform targeted reaching. Instead, SCS02 was instructed to reach from a point close to their chest and to the furthest of 3 lines that they could reach. This task was only used in the frequency optimization experiment for SCS02.

### Biomechanical arm simulation and feedback controller design

#### The PD controller

We modeled the neural control of torque generation at each joint as a pair of proportional-derivative (PD) controllers that regulate activation of the flexor and extensor muscle groups. Angular displacement is positive in the extensor direction. The input to each PD controller is the error state vector, which represents the difference between the current and target angular position and velocity states. The target positions were set to the joint angles recorded when a target was reached, and the angular position error (Δθ*_i_*) is the difference between the target joint angle (θ_*T*_) and the actual angle (θ_*i*_) at time *i*. Since the target angular velocity is 0, the angular velocity error is simply the actual angular velocity 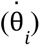. The control policy for regulating activation of each muscle group is represented by the following equations:

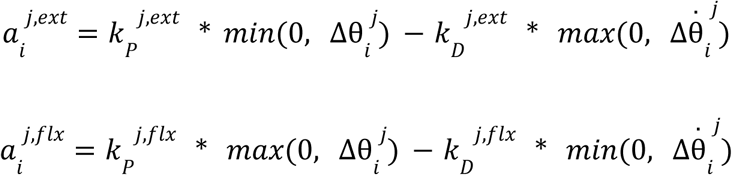

Where 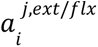 is the muscle activation signal at time *i* for the extensor (*ext*) or flexor (*flx*) muscle group at joint *j* (shoulder or elbow). The proportional (k_P_) and derivative (k_D_) gain terms are tuned for each actuator. The saturation functions ensure that the flexor/extensor torques are nonzero only when the actuator is engaged to generate torque in its respective direction. For example, 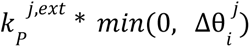 is 0 when 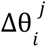 is positive, and 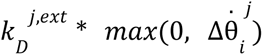 is 0 when 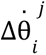 is negative.

The muscle activation signal is converted into torque through a first order transfer function (see Figure 1e for a schematic), and the flexor and extensor torques are summed for each joint to produce the total joint torque.

### Feedback Controller Optimization

The feedback controller gains were determined by Bayesian optimization with a Gaussian prior. Bayesian optimization entailed searching for the controller gains that produced torque profiles that are the closest to experimental values based on the joint angles and angular velocities measured during the experiments. Below we describe the gain search algorithm.

#### Gain Search Algorithm

Joint kinematics were measured using the Kinarm while participants performed planar reaching movements. Joint kinematics were used to estimate joint torques as described in a later subsection. The gain parameters for the PD controller model were fit using Bayesian Optimization to find a set of PD controller gains that minimized the error between the measured and simulated joint torques, as described in the following loss function:

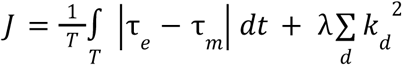

where λ controls the the penalty for selecting larger gain values, T is the duration of the trial, and *k_d_* are the individual gain values, where *d* denotes proportional/derivative flexor/extensor gains. The loss is dominated by the absolute error of the model relative to the experimental data normalized to the duration of the trial, with an additional component to penalize higher gains, ensuring that the model fits the data with the minimum value possible for each gain. The λ parameter was set to 0.001 for all model fittings reported in this paper. Gain values are initiated at random values (typically within a range of 0-10 N-m/rad and N-m-s/rad). Every new iteration, the gains were updated by adding a random number with standard deviation equal to 50% of the gain range (i.e. 5 in the case of [0, 10] range). Every time the model made an improvement, the gain search was narrowed by halving the gaussian noise. The optimization was stopped when a desired loss was met. This optimization loop was then repeated a predetermined number of times (5 unless otherwise stated) and the best gains were averaged over these iterations to account for the stochastic nature of the search. Desired loss was adjusted manually between targets and participants as to best fit each torque curve. The gain search was implemented as a custom-written script in Matlab, and is provided in the Source Code section.

#### Gain Normalization

To assess the individual torque contributions of different controller gains, we normalized the gains using the ratio of their time-integrated torque generated to the total torque produced. That is, the normalized gains, 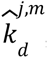, are related to the unnormalized gains, 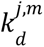, through the equation:

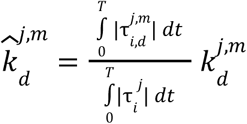

where *j* is the respective joint (shoulder or elbow), *d* denotes proportional and derivative gains for the flexor and extensor directions indicated by 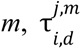 is the component torque for each unidirectional actuator, τ*_j_* denotes the total joint torque applied at time *i* at joint *j*, and *T* is the duration of the reaching trial. Unlike 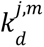, the normalized gain values, 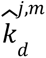 are more indicative of the relative contribution of each feedback gain to the total torque produced.

#### Experimental Torque Calculation

In the cases where torque was not recorded by the Kinarm system during the experiment, torque was instead calculated using the recorded joint angles, velocities and accelerations by using a two link manipulator as a model. By finding the theoretical kinetic energy (*T*) at any given time step and assuming potential energy (*U*) was negligible, we find that the lagrangian (*L*) is simply the kinetic energy at any given time, and joint torques (τ ) can be calculated through the Euler-Lagrange equation 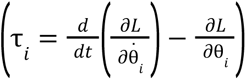 such that:

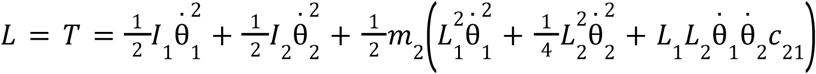

yielding:

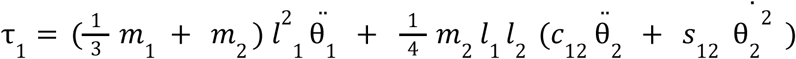

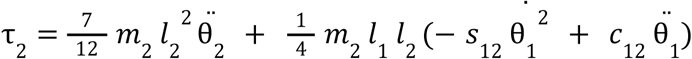

where *m*_*i*_ and *L*_*i*_ denote the masses and lengths of each arm segment, respectively (1 referring to the upper arm , while 2 refers to the forearm), θ*_i_* and 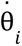 denote the angle and angular velocity of an arm segment relative to the horizontal, and *c*_12_ and *s*_12_ denote cos (θ_1_ – θ_2_) and sin (θ_1_ – θ_2_) respectively. Segment masses and lengths were estimated based on ^14^. For simplicity, each segment was treated as a thin rod with moment of inertia 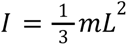 where *m* and *L* denote the segment mass and length, respectively. We did not include damping in the torque calculations for two reasons. First, reaching speeds were slow, making damping effects negligible. Second, a damping term would shift the absolute values of the derivative gain estimates but would not affect the *difference* in derivative gains between STIM OFF and STIM ON. Because these torque estimates were used solely to compare experimental control gains (and not to match the arm simulation torques), omitting damping is justified. Since minor changes in inertial properties will not affect the qualitative structure of the controller gains, we used the same inertial properties for all participants when torque was estimated (see table below for parameters).

#### Model Performance Assessment

The following metrics were used to assess the model’s fitted torque profiles in comparison to experimental values:

- **Normalized RMSE (NRMSE):** RMSE measures the quadratic residuals of the calculated (i.e. model) τ_*m*_ from the reference (i.e. experimental) signal τ_*e*_. This metric is normalized to the peak-to-peak amplitude of τ*_e_* to accommodate for the widely different ranges of torque profile amplitudes during reaching across participants, targets, and stimulation parameters. This metric was used to assess how the model deviated, on average, from the experimental values.
- **Signed Normalized Error (SNE):** We defined Signed Error as the average error between τ_*m*_ and τ*_e_* across the duration of the trial, which was likewise normalized to the peak-to-peak amplitude of τ. This metric was used to assess how well the model’s predicted torque tracked the average experimental torque profiles.

#### Two-link biomechanical model

We built a two-link biomechanical model in Matlab Simulink (MathWorks) to simulate reaching behavior under conditions where the feedback controller gains are optimized to produce smooth reaching trajectories. The arm was modeled as a two-link arm, with segments representing the upper and lower arms. The inertial and geometric parameters of the two-link model were identical to those used in experimental torque calculations (see earlier). Joints were modeled as damped revolute joints where the shoulder was fixed while the elbow could move freely in the 2D plane ^26,27^. The parameters used in the model are summarized in the table below, and the model simulation files are included in the Source Code section. We used the same optimization model to search for gain structures that are optimal for 2D reaching using the two-link model. The class of objective functions used is represented by

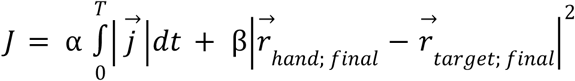

Where 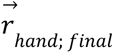 and 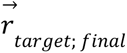 represent the final position of the hand and the target in 2D plane during the last, respectively. α and β are free parameters to assign different weights for reaching the target and generating smooth trajectories, respectively. T denotes the total simulation time, set typically to 5 seconds. Adjusting T forces the model to choose gains that result in faster reaching times. 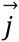 denotes the jerk vector of the hand, or 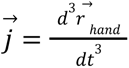. Jerk is a measurement of how smooth trajectories are; lower jerk indicates smooth reaching.

| Symbol | Value | Description |
| --- | --- | --- |
| $\alpha$ | $10^{-6}$ | Coefficient of hand jerk in the objective function |
| $\beta$ | 10 | Coefficient of deviation from target term in the objective function |
| $L_{UA}$ | 0.35 m | Length of upper arm segment |
| $L_{LA}$ | 0.35 m | Length of lower arm segment |
| $m_{UA}$ | 2.8 kg | Mass of upper arm segment |
| $m_{LA}$ | 2.1 kg | Mass of lower arm segment |
| $c$ | 1.5 N*m/(rad/s) | Joint damping coefficient |

#### Performance metrics for planar reaching task

To assess performance at the kinematics level, we used multiple metrics in this work:

##### Reach Time

The time it takes to reach the designated target after receiving a ‘go’ cue from the KinArm system.

##### Path Efficiency

The performance of each reaching path in comparison to the ideal path such that:

- Path Traveled 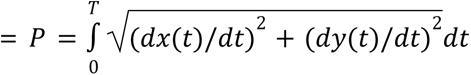
- Path Efficiency = *D*/*P*

where T denotes the total reach time, x and y denote hand positions, dx/dt and dy/dt denote the hand velocities measured during reaching, and D denotes the straight line distance from start to target. This measure is used as it scales between 0 and 1 and can be interpreted as a percentage of the participant’s performance relative to the best possible performance (100% being a straight line).

##### Number of Elbow Velocity Peaks

The number of peaks in the speed profile of the elbow during the reaching phase of the task. Fewer peaks is associated with smoother trajectories.

##### Reaching Success Rate

The total number of successful trials (i.e. where the participant made it to the target in under 20 seconds) divided by the total number of trials. Success rates were calculated using all experimental trials included for each participant

### Figure Generation

Plots were generated in either Matlab (MathWorks) or Python, and were then styled and exported into vector graphics format using InkScape and BioRender.

### Statistics

We used paired t-tests to compare performance metrics and model fit gains for STIM OFF versus STIM ON for the same participant, and unpaired t-tests for other comparisons (e.g. comparisons between different participants, etc.). All samples were tested using the Shapiro-Wilk test prior to using t-test to confirm the normality of sample distributions. When appropriate, for multiple t-tests of groups of conditions related to one another, we used multiple t-test corrections using the Benjamini Hochberg method for false discovery rate correction. We further used Cohen’s d-values to assess differences between samples when appropriate. We used a cutoff level for statistical significance of 0.05 throughout the study.

## Data availability

Data used to generate the figures of the paper is provided in the Source Data section. Access to raw data may be granted if a reasonable request is submitted to D.J.W. at.

## Source Data

Movement Metrics (Figures 2 and 6, Extended Data Table 1)

NE and RMSE (Figure 3): See Extended Data Table 2.

Model Fit Gains (Figure 3-6, Extended Data Figures 1, 2 and 4, Supplementary Figure 3)

## Code availability

The code used to generate model fits to experimental data, and the two-link arm Simulink simulation files are all publicly available in the GitHub repository at link.

## Acknowledgements

We would like to acknowledge Hazel Cline, Elle Brough, and Cemal Ozis for their contributions during the exploratory phase of this study. The experimental parts of this study were executed through the support of National Institutes of Health Brain Initiative grant no. UG3NS123135-01A1 to M.C. and D.J.W. We would like to thank Serhii Bahdasariants for valuable feedback on the preprint. O.R.’s time during manuscript revisions was supported by the Lundbeck Foundation Postdoctoral Fellowship.

## Author information

These authors contributed equally: Omar Refy, Luigi Borda, and Jacob Hsu

## Contributions

Model conceptualization: O.R., L.B., and D.J.W.

Model development: O.R.

Model testing and validation: O.R., J.H. and L.B.

Model revisions: O.R., D.J.W., H.G.

Experimental setup conceptualization and design: G.W., E.P., L.F., J.K., M.C. and D.J.W.

Experimental data collection: L.B., N.V., E.C., R.F. and M.P.

Experimental data analysis: J.H., L.B. and O.R.

Surgical procedures: P.G.

Motor and behavioral assessments: A.B.

Drafted manuscript: O.R., J.H., L.B., and D.J.W.

Manuscript revision and editing: O.R., and D.J.W.

Manuscript approval: D.J.W.

## Ethics declarations

D.J.W., M.C., M.P. and P.C.G. are founders and shareholders of Reach Neuro, a company developing spinal cord stimulation technologies for stroke. E.P. has interest in Reach Neuro due to personal relationship with M.C. All other authors declare no competing interests.

## Extended data

**Extended Data Table 1.** Percent changes in reaching performance metrics in the STIM ON versus STIM OFF conditions. (*): p < 0.05, (**): p < 0.01, (***): p < 0.001.

| Subject | Baseline FM-UEA [0-66] | Baseline Success Rate (%) | Path Efficiency | Reach Time | Elbow Velocity Peaks |
| --- | --- | --- | --- | --- | --- |
| SCS02 | 15 | 20 (80) | 160.00% (***) | 56.3% (*) | 140.00% (**) |
| SCS03 | 28 | 0 (100) | 1.25% (n.s.) | 6.6% (n.s.) | 6.98% (n.s.) |
| SCS04 | 23 | 5 (83) | 25.93% (*) | 29.4% (n.s.) | 75.00% (**) |
| SCS07 | 34 | 0 (100) | 4.48% (n.s.) | 23.6% (***) | 11.54% (n.s.) |
| SCS08 | 32 | 0 (100) | -0.69% (n.s.) | 9.4% (n.s.) | 9.4% (n.s.) |

**Extended Data Table 2.** Summary of reaching performance metrics of 3 control and 4 stroke participants (coded HC0X and SCS0X, respectively). Participants in the stroke group completed the task with both limbs, and data for the non-paretic limb is included in the table. Metrics are for the reaching out phase. Values are reported as mean (standard deviation).

| Participant | Condition | Number of Trials | Reach Time (s) | Path Efficiency | # Elbow Velocity Peaks | Success Rate (%) |
| --- | --- | --- | --- | --- | --- | --- |
| HC01 | Healthy | 3 | 1.86 (0.22) | 0.99 (0.01) | 1.7 (0.6) | 100 |
| HC02 | Healthy | 3 | 1.85 (0.20) | 0.93 (0.01) | 2.3 (0.6) | 100 |
| HC03 | Healthy | 3 | 2.10 (0.37) | 0.95 (0.03) | 1.7 (0.6) | 100 |
| HC04 | Healthy | 3 | 2.08 (0.16) | 0.96 (0.00) | 3.3 (0.6) | 100 |
| HC05 | Healthy | 3 | 2.44 (0.28) | 0.98 (0.01) | 3.0 (1.0) | 100 |
| HC06 | Healthy | 3 | 2.95 (0.34) | 0.98 (0.01) | 5.3 (2.1) | 100 |
| HC07 | Healthy | 3 | 2.05 (0.20) | 0.97 (0.01) | 2.0 (1.0) | 100 |
| HC08 | Healthy | 3 | 1.76 (0.07) | 0.92 (0.01) | 2.0 (0.0) | 100 |
| HC09 | Healthy | 3 | 2.10 (0.42) | 0.96 (0.02) | 3.0 (1.0) | 100 |
| HC10 | Healthy | 3 | 2.15 (0.23) | 0.97 (0.01) | 1.7 (0.6) | 100 |
| HC11 | Healthy | 3 | 2.00 (0.07) | 0.97 (0.02) | 2.3 (0.6) | 100 |
| HC12 | Healthy | 3 | 2.32 (0.08) | 0.96 (0.01) | 2.3 (0.6) | 100 |
| SCS02 | STIM OFF | 30 | 7.05 (4.70) | 0.20 (0.21) | 11.6 (10.7) | 80 |
| SCS02 | STIM ON | 30 | 4.51 (4.42) | 0.52 (0.23) | 5.1 (7.9) | 100 |
| SCS03 | STIM OFF | 30 | 3.22 (1.10) | 0.80 (0.09) | 4.6 (2.7) | 100 |
| SCS03 | STIM ON | 30 | 3.02 (1.20) | 0.81 (0.09) | 4.3 (2.8) | 100 |
| SCS03 | Non-paretic limb | 14 | 1.46 (0.44) | 0.90 (0.05) | 3.3 (1.7) | 100 |
| SCS04 | STIM OFF | 24 | 4.17 (2.00) | 0.54 (0.19) | 6.5 (3.7) | 83 |
| SCS04 | STIM ON | 18 | 3.23 (2.00) | 0.68 (0.12) | 3.7 (1.3) | 88 |
| SCS04 | Non-paretic | 15 | 1.42 (0.42) | 0.91 (0.05) | 5.2 (1.2) | 100 |
|  | limb |  |  |  |  |  |
| SCS07 | STIM OFF | 60 | 2.04 (0.84) | 0.65 (0.10) | 2.9 (1.6) | 100 |
| SCS07 | STIM ON | 45 | 1.65 (0.64) | 0.68 (0.14) | 2.6 (1.6) | 100 |
| SCS07 | Non-paretic limb | 15 | 1.20 (0.37) | 0.90 (0.07) | 2.4 (1.4) | 100 |
| SCS08 | STIM OFF | 30 | 2.26 (1.22) | 0.74 (0.10) | 5.2 (3.3) | 100 |
| SCS08 | STIM ON | 18 | 2.04 (0.78) | 0.74 (0.14) | 4.8 (2.4) | 100 |
| SCS08 | Non-paretic limb | 90 | 1.03 (0.31) | 0.95 (0.03) | 1.6 (0.5) | 100 |

**Extended Data Figure 1.**
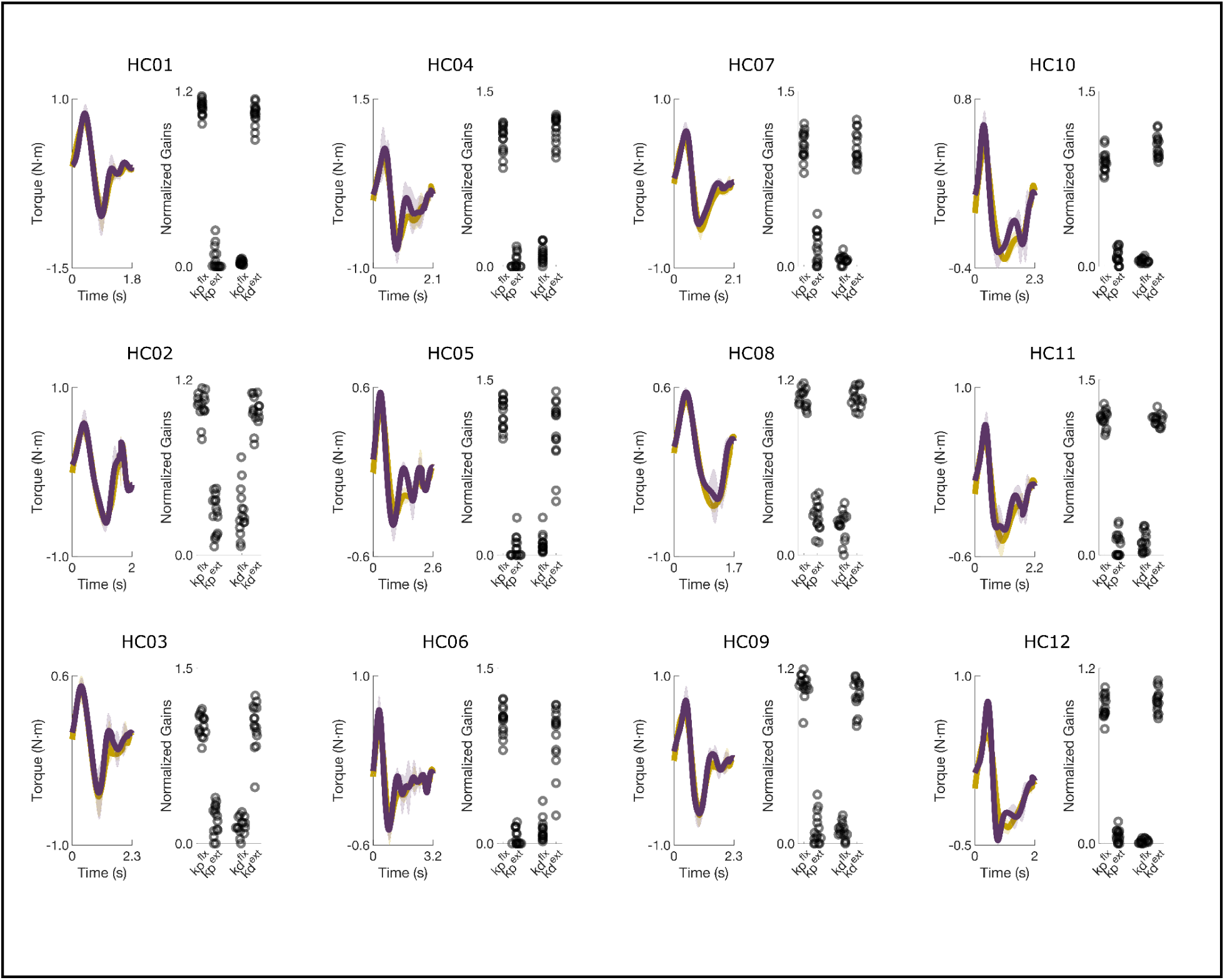
Measured and simulated torque profiles for the shoulder during planar reaching performed by healthy control participants. The scatterplots show the normalized controller gains for the PD controller simulations that were fit to the data. The controller gains exhibit similar U-shape and inverted U-shape patterns across subjects for the elbow and shoulder, respectively. Corresponding NRMSE and SNE values are reported in Extended Data Table 2.

**Extended Data Figure 2.**
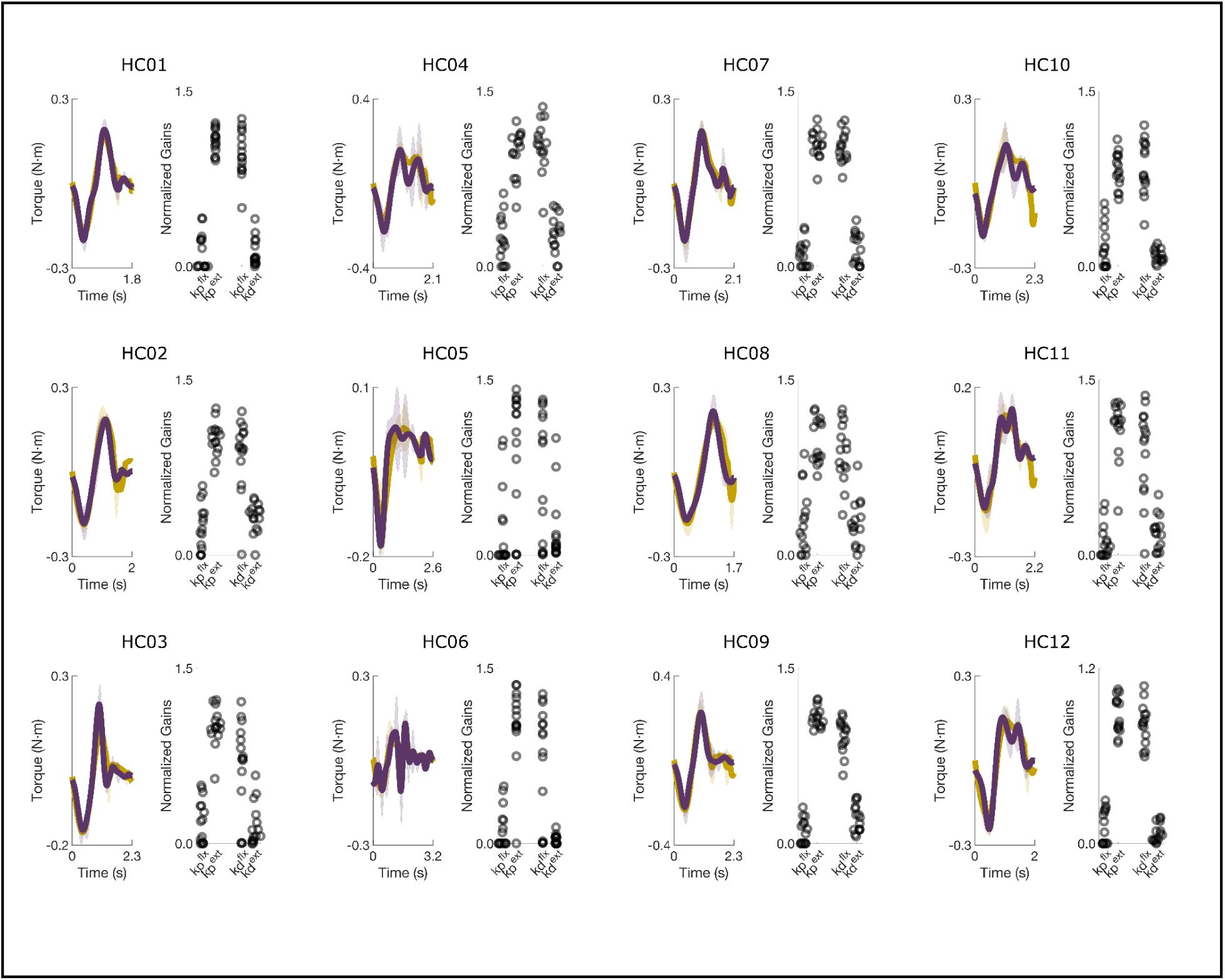
Measured and simulated torque profiles for the elbow during planar reaching performed by healthy control participants. Values represent normalized gains. Data are of the same trials as Extended Data Figure 1. Corresponding NRMSE and SNE scores are reported in Extended Data Table 2.

**Extended Data Figure 3.**
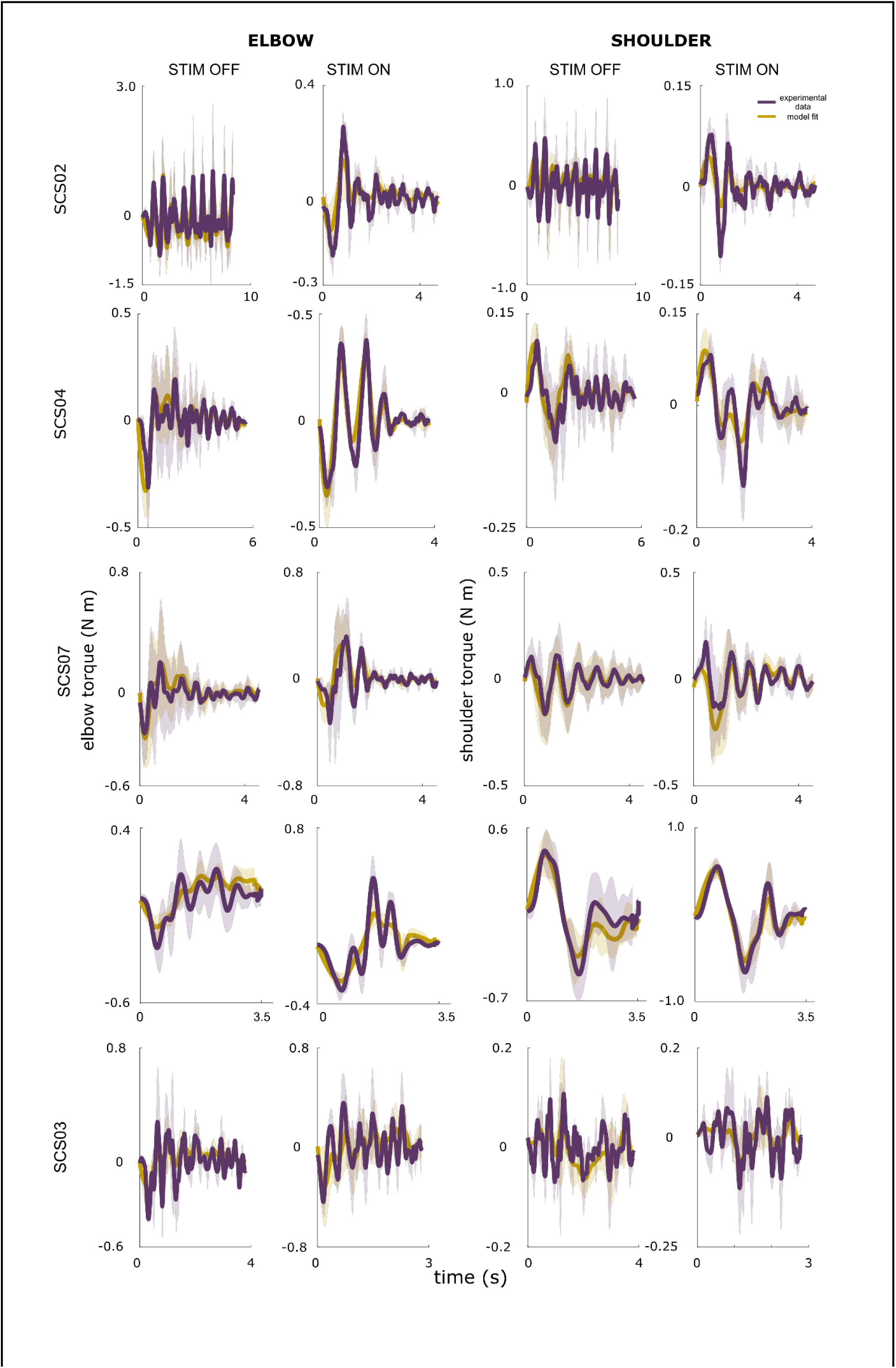
Measured and simulated torque profiles for the shoulder and elbow during planar reaching performed by five participants in the stroke group. The model produced good fits for both elbow and shoulder for all participants under both STIM OFF and STIM ON conditions. Corresponding NRMSE and SNE scores are reported in Extended Data Table 2.

**Extended Data Figure 4.**
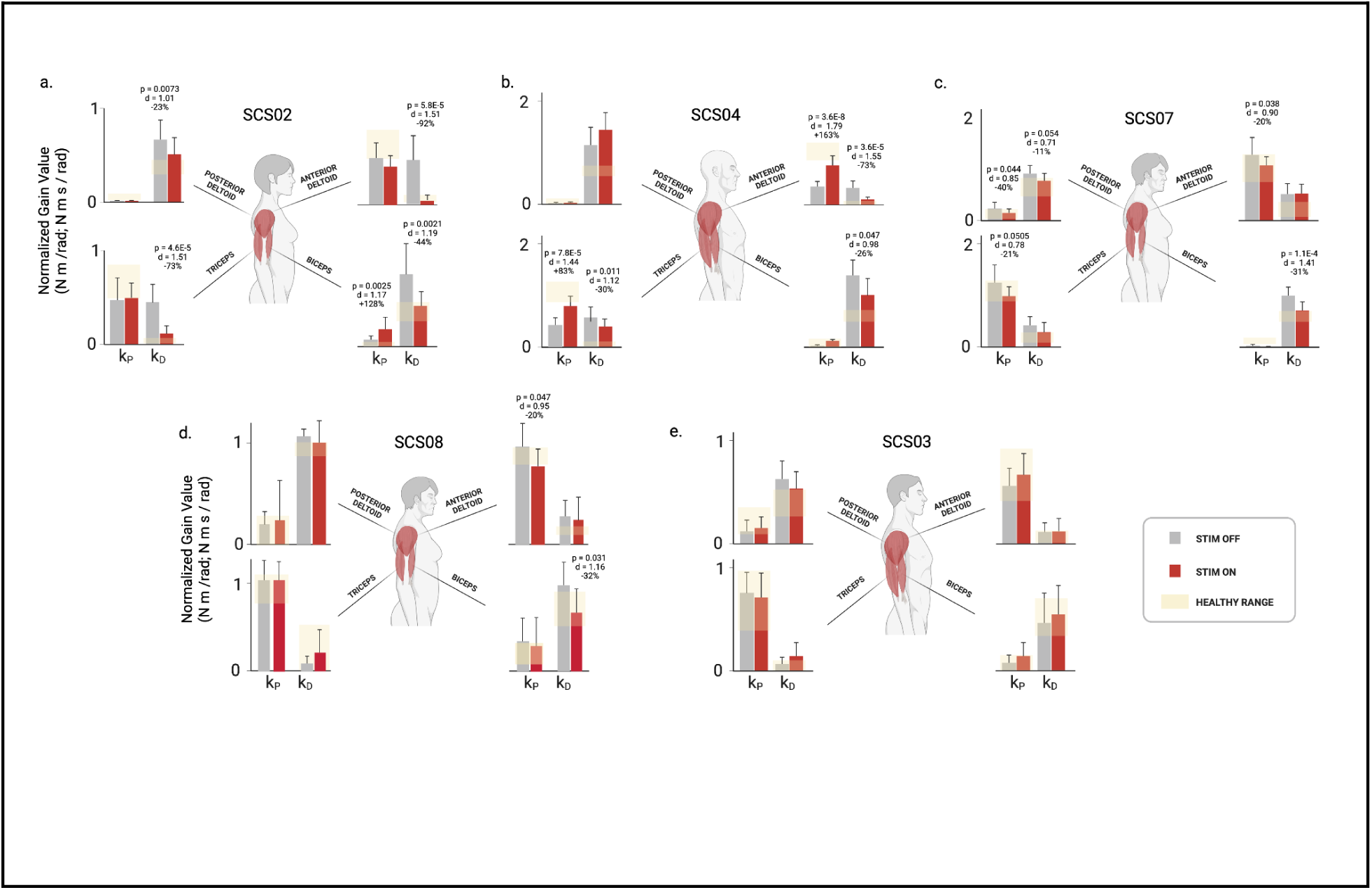
Controller gains for the shoulder and elbow for the five participants in the stroke group. The controller gains are the result of fitting the PD controller simulation to the reaching data for the center-out task presented in Extended Data Figure 2. SCS02 (panel a) showed the greatest improvements in reaching performance metrics (Table 1) in the STIM ON condition, and the underlying changes in control gains provide insights into why. Looking at the elbow joint, we observed that the derivative gains of both elbow flexors and extensors were significantly reduced under STIM ON. In particular, the elbow flexor derivative derivative gain was reduced by 44% (p = 0.0021, d = 1.19), and the extensor derivative gain was reduced by 73% (p < 0.001, d = 1.51). Both derivative gains of the shoulder were also reduced. The derivative gains of the shoulder flexor and extensor were reduced by 92% (p < 0.001, d = 1.51) and 23% (p = 0.007, d = 1.01), respectively. Notice that all gains are tuned towards healthy ranges, with the exception of the flexor proportional gain, which increased with STIM ON. SCS04 (panel b) also showed significant reduction in 3 out of 4 derivative gains (albeit less drastic due to a better baseline compared to SCS02). In particular, the derivative gains of the elbow flexor and extensor were reduced by 26% (p = 0.047, d = 0.98) and 30% (p = 0.01, d = 1.12), respectively, while that of the shoulder flexor was reduced by 73% (p < 0.001, d = 1.55). In addition, the proportional gains of both the elbow extensor and the shoulder flexor (i.e. the two proportional ballistic gains, which act to drive the joints towards the target during the ballistic phase) increased by 83% (p < 0.001, d = 1.44) and 163% (p < 0.001, d = 1.79, respectively. Similar to SCS02, tuning of the control gains in SCS04 were towards healthy ranges, but spanned both proportional and derivative gains. SCS07 (panel c) showed changes that were less pronounced compared to both SCS04 and SCS02. Two out of 4 derivative gains showed decrease closer to healthy ranges, namely the shoulder extensor and the elbow flexor. The shoulder extensor derivative gain showed a trend of 11% reduction (p = 0.054, d = 0.71), while the elbow flexor showed a significant 31% reduction (p < 0.001, d = 1.41). Contrary to SCS04 who had proportional ballistic gains lower than healthy ranges, SCS07 had higher baseline compared to healthy ranges, which was reduced by SCS. In particular, the elbow extensor proportional gain showed a trend of 21% reduction (p = 0.051, d = 0.78), and the shoulder flexor showed a significant 20% reduction (p = 0.038, d = 0.90). Overall, SCS07 showed the same trend of control gains tuned closer to healthy control ranges (albeit in a slightly different way compared to SCS04 and SCS02), as summarized in panel c. SCS03 showed no significant difference between all gains, which is consistent with his close to healthy baseline reaching, and hence the minimal effects of SCS. Overall, 3 out of 4 participants showed control gain tuning closer to healthy ranges (with the exception of SCS03 (panel d) who had baseline close to healthy to begin with). Notably, while different participants showed gain tuning closer to healthy in different ways. SCS02 showed overall reduction in derivative gains, SCS04 showed reduction in derivative gains and increase in proportional gains, while SCS07 showed reduction in derivative gains and proportional gains alike. The tuning of the gains was individualized, and depended on the participants’ baseline gains. This highlights both that gain tuning is not arbitrary, and the individualized nature of the effects of neuromodulation on the neural controller of the arm.

**Extended Data Figure 5.**
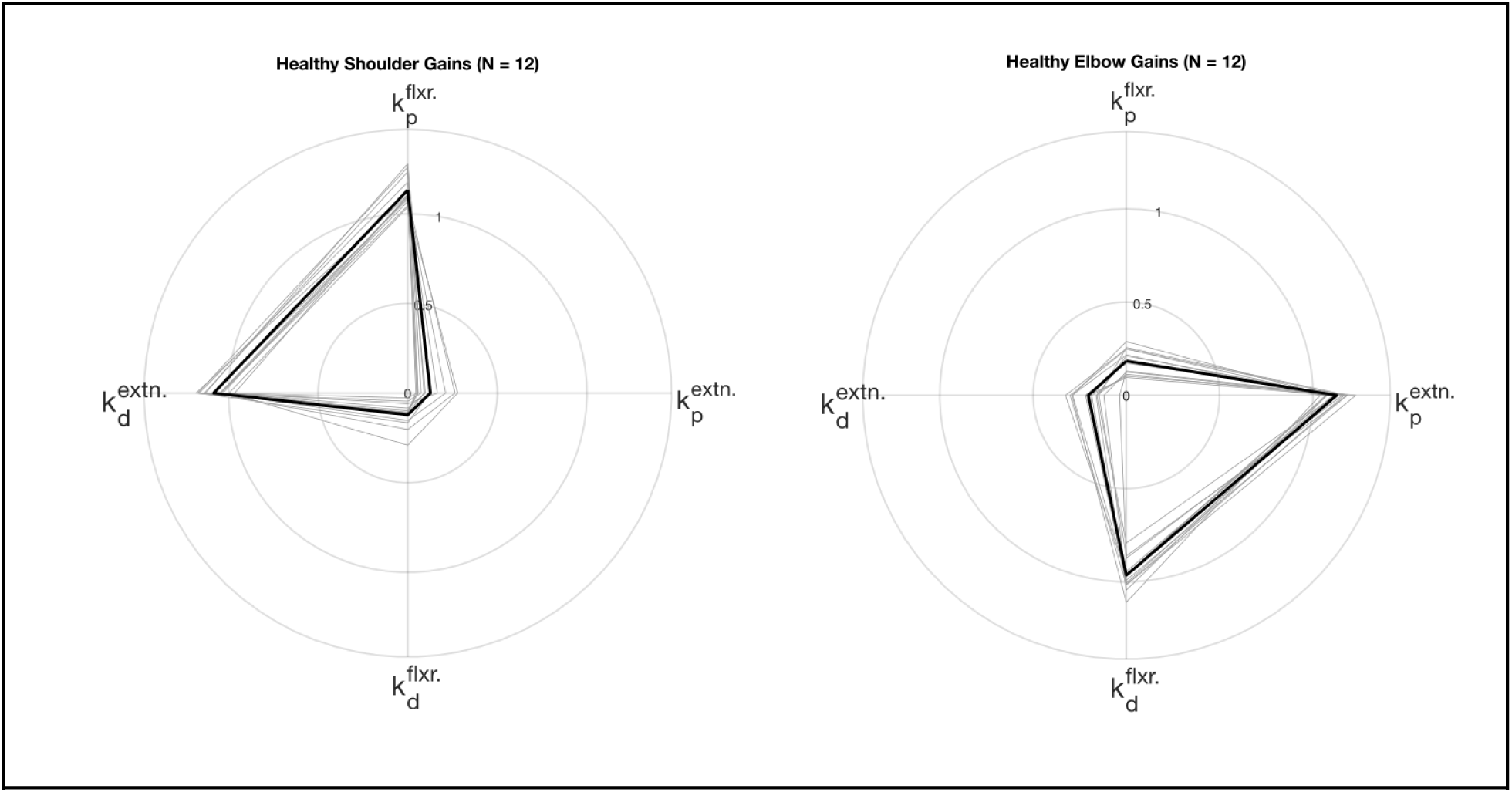
Average gain fits for all healthy participants (N = 12 participants; n = 3 trials each, 5 model fits for each trial) for elbow and shoulder joint during forward reaching. Individual participant values are shown in thin gray and group average is shown in thicker black lines.

**Extended Data Table 3.**
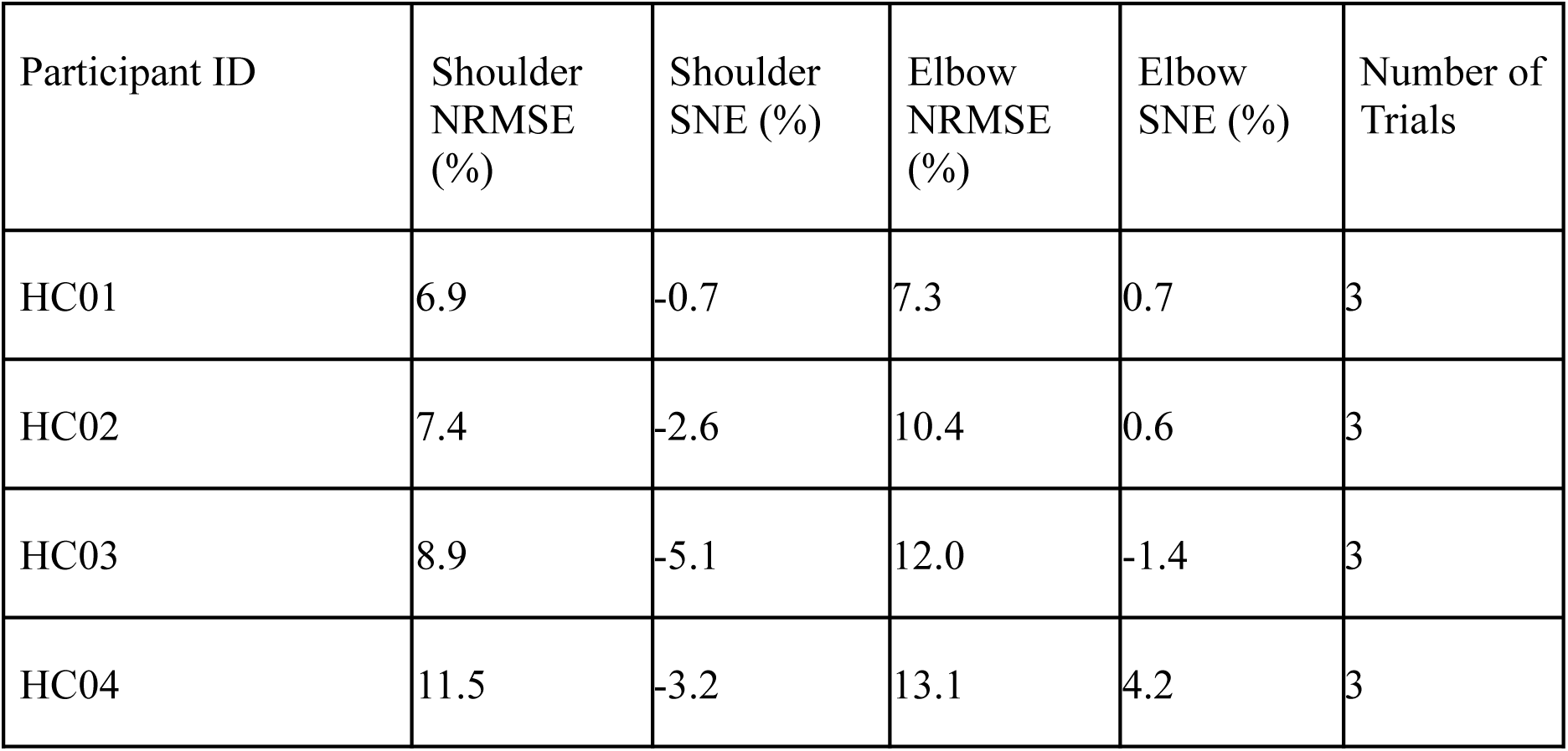
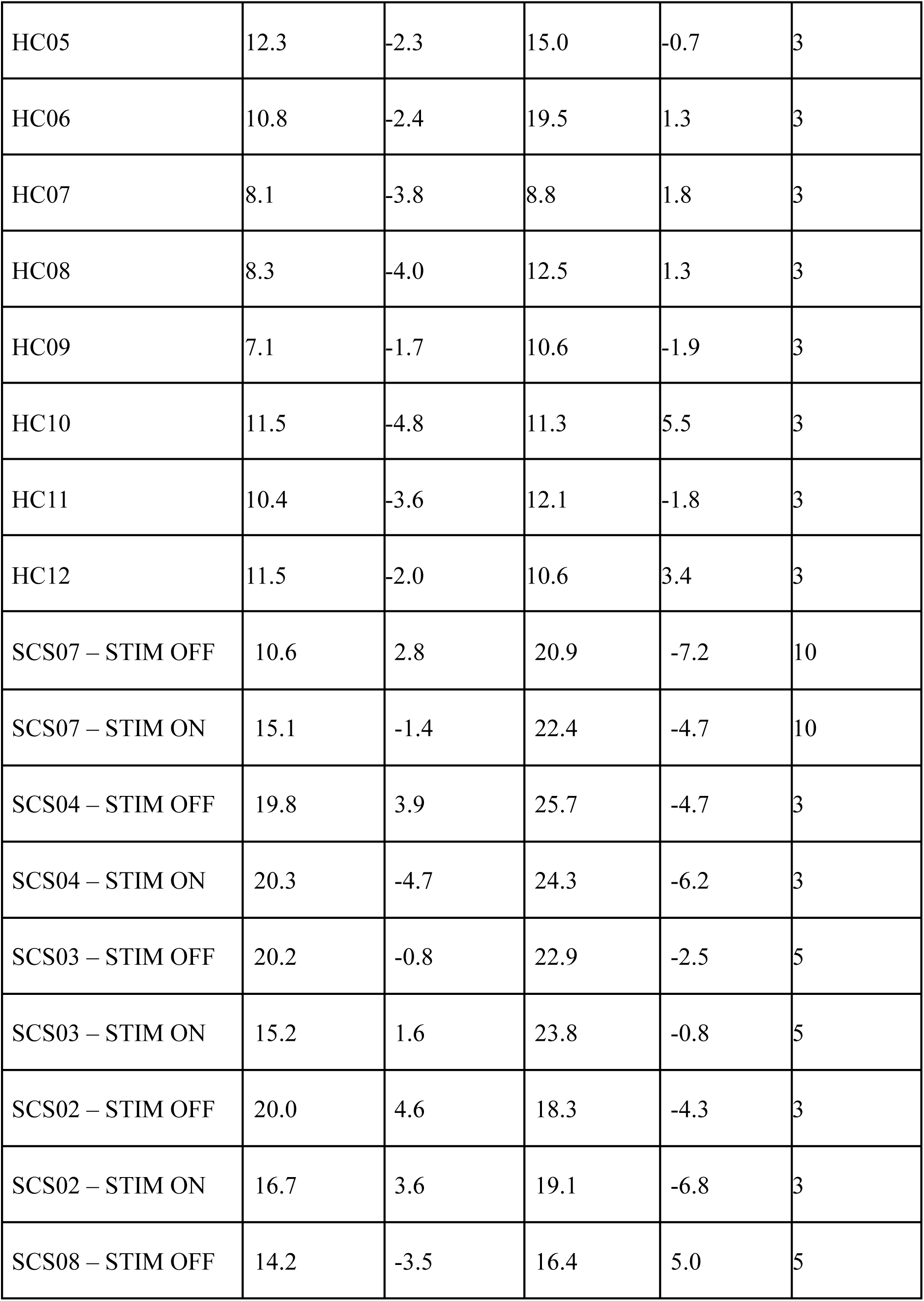
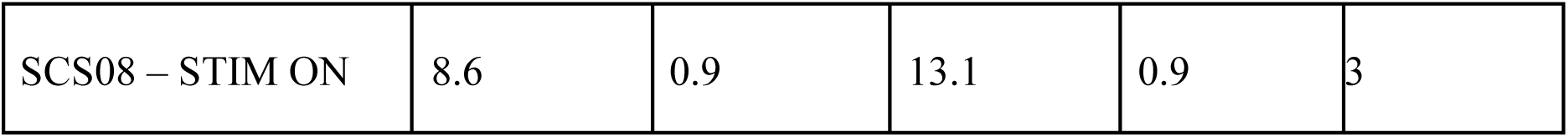
NRMSE and SNE values for all participant model fits. Values are the averages of 5 model fits per trial for all participants.

| Participant ID | Shoulder NRMSE (%) | Shoulder SNE (%) | Elbow NRMSE (%) | Elbow SNE (%) | Number of Trials |
| --- | --- | --- | --- | --- | --- |
| HC01 | 6.9 | -0.7 | 7.3 | 0.7 | 3 |
| HC02 | 7.4 | -2.6 | 10.4 | 0.6 | 3 |
| HC03 | 8.9 | -5.1 | 12.0 | -1.4 | 3 |
| HC04 | 11.5 | -3.2 | 13.1 | 4.2 | 3 |
| HC05 | 12.3 | -2.3 | 15.0 | -0.7 | 3 |
| HC06 | 10.8 | -2.4 | 19.5 | 1.3 | 3 |
| HC07 | 8.1 | -3.8 | 8.8 | 1.8 | 3 |
| HC08 | 8.3 | -4.0 | 12.5 | 1.3 | 3 |
| HC09 | 7.1 | -1.7 | 10.6 | -1.9 | 3 |
| HC10 | 11.5 | -4.8 | 11.3 | 5.5 | 3 |
| HC11 | 10.4 | -3.6 | 12.1 | -1.8 | 3 |
| HC12 | 11.5 | -2.0 | 10.6 | 3.4 | 3 |
| SCS07 – STIM OFF | 10.6 | 2.8 | 20.9 | -7.2 | 10 |
| SCS07 – STIM ON | 15.1 | -1.4 | 22.4 | -4.7 | 10 |
| SCS04 – STIM OFF | 19.8 | 3.9 | 25.7 | -4.7 | 3 |
| SCS04 – STIM ON | 20.3 | -4.7 | 24.3 | -6.2 | 3 |
| SCS03 – STIM OFF | 20.2 | -0.8 | 22.9 | -2.5 | 5 |
| SCS03 – STIM ON | 15.2 | 1.6 | 23.8 | -0.8 | 5 |
| SCS02 – STIM OFF | 20.0 | 4.6 | 18.3 | -4.3 | 3 |
| SCS02 – STIM ON | 16.7 | 3.6 | 19.1 | -6.8 | 3 |
| SCS08 – STIM OFF | 14.2 | -3.5 | 16.4 | 5.0 | 5 |
| SCS08 – STIM ON | 8.6 | 0.9 | 13.1 | 0.9 | 3 |

## Supplementary information

### Interaction Torque Contributions and Choice of Control Gain Structure

For single joint movements, torque is related to angular acceleration simply by a single factor (i.e. moment of inertia of the arm segment moving). In two-joint arm movements, movement of one joint can generate torque at the other (i.e. interaction torques). Interaction torques are typically more prominent at some arm configurations over others (e.g. fully straight arm versus elbow and shoulder slightly flexed), and at higher angular speeds of either joint, as can be inferred from the equations of motion. In all the planar movement trials we performed, due to relatively lower speeds and ranges of arm configurations, torques of the elbow and the shoulder were mostly dominated by torque generated at individual joints. That is, interaction torques were quite minimal, especially for stroke participants due to their lower reaching speeds. This justifies the choice of control model where each joint is controlled independently from the other (i.e. the elbow PD controller only generates torque based on elbow feedback, and same for the shoulder). Plots of example interaction and generated torques for shoulder and elbow joints of one healthy and two stroke participants are shown below to illustrate the idea.

**Supplementary Figure 1.**
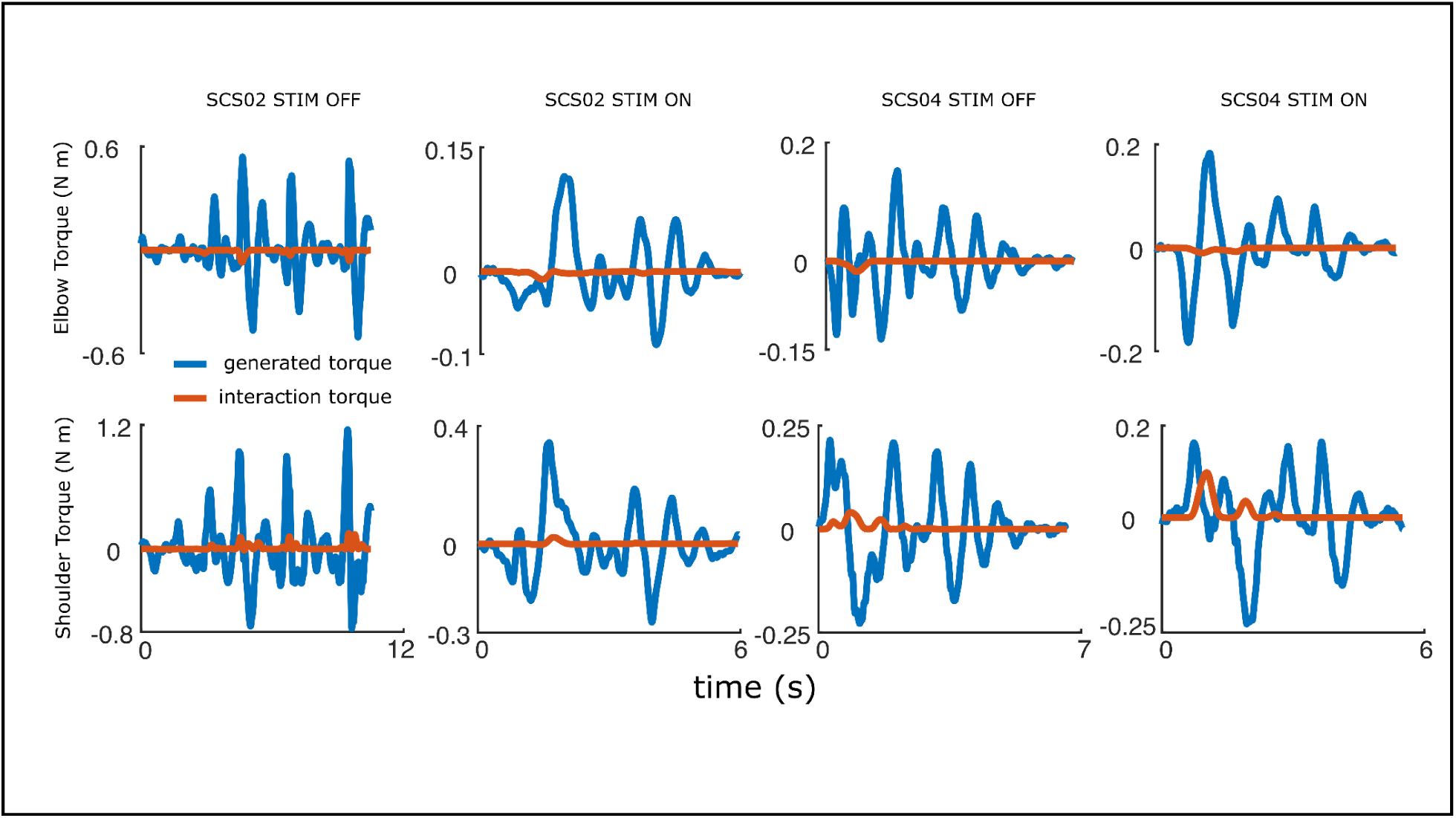
Interaction torques were often not very prominent in the task we were modeling, likely due to small angle ranges and relatively slow reaching speeds.

### Non-paretic arm versus healthy model fits

**Supplementary Figure 2.**
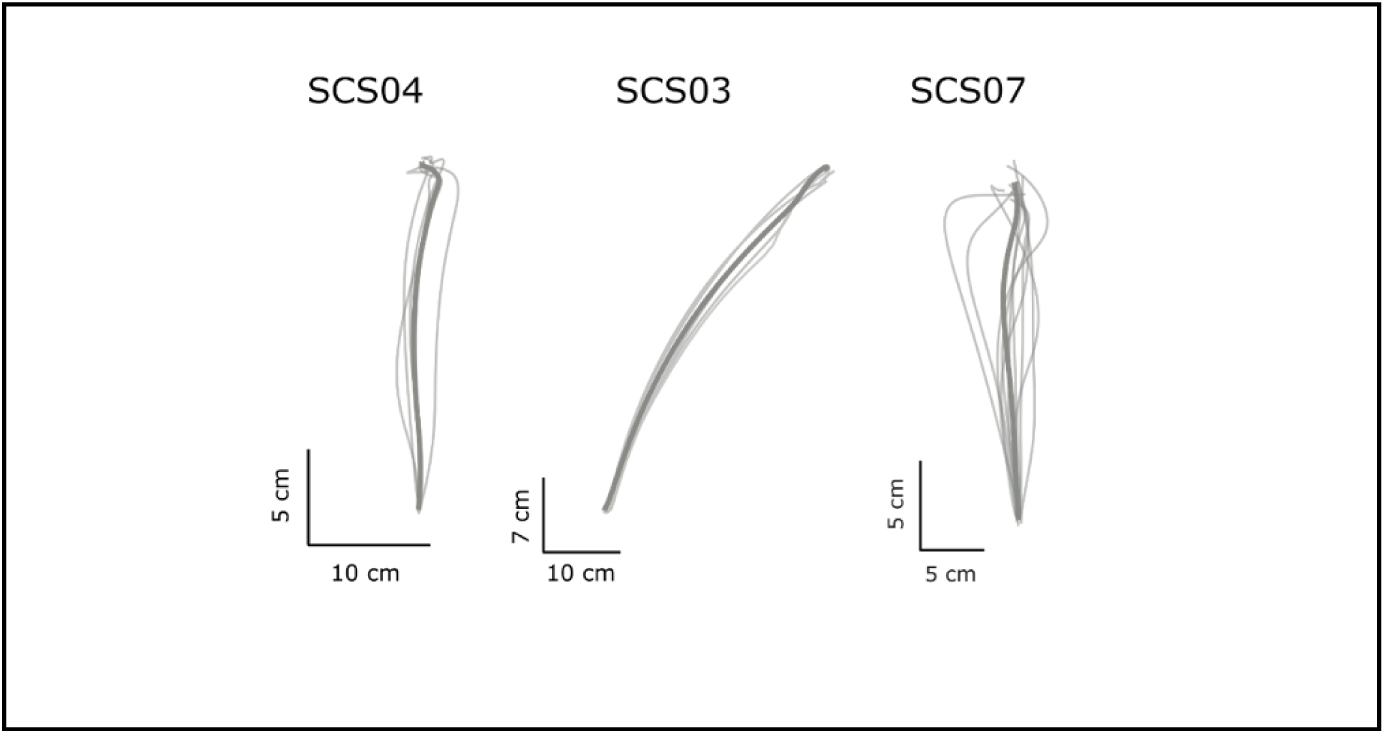
Non-paretic arm showed closer to healthy point-to-point reaching. Non-paretic arm reaching was not recorded during planar arm reaching for SCS02.

## Supplementary Videos

Supplementary Video 1: HC09 component torques

Supplementary Video 2: SCS02 STIM OFF component torques

Supplementary Video 3: SCS02 STIM ON component torques

Supplementary Video 4: Simulated arm with control structure tuned to produce smooth reaching.

Supplementary Video 5: Simulated arm with suboptimal control structure (increased flexor derivative gain).

## Notes

### Clinical Trial

NCT04512690

### Author Declarations

This study was conducted as a substudy under the Spinal Cord Stimulation for Restoration of Arm and Hand Function in People With Subcortical Stroke (NCT04512690) trial aimed at evaluating the effects of spinal cord stimulation for restoring arm and hand function post-stroke. All experiments were approved by the University of Pittsburgh Institutional Review Board, and were conducted in accordance with the Declaration of Helsinki. All participants provided written informed consent.

### Summary of Updates

This revision is for fine tuning of narrative in the abstract, introduction and discussion section. Results, figures and main conclusions remain unchanged.

